# What innovations (including return to practice) would help attract, recruit, or retain NHS clinical staff? A rapid evidence map

**DOI:** 10.1101/2022.05.10.22274894

**Authors:** Deborah Edwards, Judit Csontos, Elizabeth Gillen, Judith Carrier, Ruth Lewis, Alison Cooper, Micaela Gal, Rebecca-Jane Law, Jane Greenwell, Adrian Edwards

## Abstract

National Health Service (NHS) waiting times have significantly increased over the past couple of years, particularly since the emergence of COVID-19. The NHS is currently experiencing an acute workforce shortage, which hampers the ability to deal with increasing waiting times and clearing the backlog resulting from the pandemic. Plans to increase the workforce, by recruiting new staff, retaining the existing NHS clinical workforce, and making return to clinical practice more attractive will require a number of approaches. This Rapid Evidence Map aimed to describe the extent and nature of the available evidence base for innovations (including return to practice) that could help attract, recruit, or retain NHS clinical staff, in order to identify the priorities and actions for a rapid review. Three options were proposed for a subsequent focused Rapid Review and discussed with stakeholders: (1) review of primary studies that have evaluated return to practice schemes; (2) review of reviews of factors that influence retention; (3) review of reviews of interventions for supporting recruitment and retention. A decision was made that option 3 would be useful to inform practice and a rapid review will be undertaken.

**Funding statement:** The Wales Centre for Evidence Based Care was funded for this work by the Wales Covid-19 Evidence Centre, itself funded by Health & Care Research Wales on behalf of Welsh Government.

**TOPLINE SUMMARY:** *What are Rapid Evidence Maps (REMs)?:* Our Rapid Evidence Maps (REMs) use **abbreviated systematic mapping or scoping review methods** to provide a description of the nature, characteristics and volume of the available evidence for a particular policy domain or research question. They are mainly based on the assessment of abstracts and incorporate an a-priori protocol, systematic search, screening, and minimal data extraction. They may sometimes include critical appraisal, but no evidence synthesis is conducted. Priority is given, where feasible, to studies representing robust evidence synthesis. They are designed and used primarily to **identify a substantial focus for a rapid review, and key research gaps in the evidence-base**. (*N.B. scoping reviews are not suitable to support evidence-informed policy development, as they do not include a synthesis of the results*.) This report is linked to a subsequent focused rapid review published as: RR00028. Wales COVID-19 Evidence Centre. A rapid review of the effectiveness of interventions/innovations relevant to the Welsh NHS context to support recruitment and retention of clinical staff. April 2022. The rapid review report is available in the WCEC library: https://healthandcareresearchwales.org/about-research-community/wales-covid-19-evidence-centre

*Background / Aim of the Rapid Evidence Map:* National Health Service (NHS) waiting times have significantly increased over the past couple of years, particularly since the emergence of COVID-19. The NHS is currently experiencing an **acute workforce shortage,** which hampers the ability to deal **with increasing waiting times** and clearing the **backlog resulting from the pandemic**. Plans to increase the workforce, by **recruiting new staff**, **retaining the existing NHS clinical workforce**, and making **return to clinical practice** more attractive will require a number of approaches. This REM aimed to **describe the extent and nature of the available evidence base for innovations (including return to practice) that could help attract, recruit, or retain NHS clinical staff**, in order to identify the priorities and actions for a rapid review.

*Key Findings:* *Extent of the evidence base*

▪ systematic or rapid reviews, 11 narrative reviews, 7 scoping reviews, 5 reviews of existing reviews (umbrella reviews), 18 primary studies, and 5 organisational reports or websites were included.
▪ evidence was categorised by the phenomena of interest: return to practice; factors influencing recruitment and/or retention; and interventions or strategies for improving recruitment and retention
▪ evidence was organised for each of the different clinical staff groups working within the NHS: nurses and midwives; doctors, including general practitioners (GPs); dentists; allied health professionals; and mixed groups of health professionals.

*Return to practice:* *Extent of the available evidence*
There was limited secondary evidence available for return to practice, and further searches for primary studies were conducted.

▪ **For nurses and midwives**, evidence was available from scoping (n=1) and systematic (n=1) reviews, primary studies (n=4), and an organisational report.
▪ **For doctors** (including GPs), evidence was available from systematic (n=2) and narrative (n=1) reviews, organisational websites (n=2), and primary studies (n=7).
▪ There was no available evidence on return to practice for **dentists**.
▪ For **allied health professionals**, evidence was available from primary studies (n=5) and an organisational website.
▪ For **mixed groups of healthcare professionals**, evidence was available from a systematic (n=1), a primary study, and an organisational report. *Summary content*
Return to practice was investigated after leaving for a variety of reasons such as mental and physical health issues, disability, maternity leave, caring responsibilities, personal or professional development opportunities, career break, moving sectors, and retirement. The evidence highlights the challenges surrounding healthcare professionals wishing to return to practice particularly with regard to “skills fade”. There are a number of routes for healthcare professionals to return to practice or training, which are well documented. However, data on their effectiveness are limited.

*Factors influencing attraction, recruitment and retention:* *Extent of the available evidence*

▪ For **nurses and midwives**, evidence was available from systematic (n=11 nurses, n=1 midwives), narrative (n=2 nurses) and scoping (n=1 nurses) reviews.
▪ For **doctors** (including GPs), evidence was available from scoping (n=2), systematic (n=5), rapid (n=1), narrative (n=1), and umbrella (n=1) reviews.
▪ There was no available evidence on factors influencing attraction, recruitment, and retention of **dentists.**
▪ For **allied health professionals**, evidence was available from systematic (n=3) and narrative (n=1) reviews.
▪ For **mixed groups of healthcare professionals**, evidence was available from systematic (n=3), rapid (n=1), and umbrella (n=1) reviews. *Summary content*
These reviews mainly focused on rural and remote areas. A broad range of factors was identified, and it has been suggested that strategies to improve attraction, recruitment and retention need to be multifaceted.

*Interventions for improving attraction, recruitment and retention:* *Extent of the available evidence*

▪ **For nurses,** evidence was available from umbrella (n=2), narrative (n=2), and scoping (n=1) reviews; no reviews were identified for **midwives**.
▪ For **doctors** (including GPs), evidence was available from scoping (n=3), systematic (n=4), and narrative (n=1) reviews.
▪ For **dentists**, evidence was available from one systematic review.
▪ For **allied health professionals**, evidence was available from a scoping (n=1) and narrative (n=1) review.
▪ For **mixed groups of healthcare professionals**, evidence was available from scoping (n=1), systematic (n=1), rapid (n=1), narrative (n=3) and umbrella (n=1) reviews.

*Summary content* Many of these reviews focused on rural and remote areas

*Implications for a Rapid Review:* Three options were proposed for a subsequent focused Rapid Review and discussed at a stakeholder meeting (held on 2^nd^ February 2022): (1) review of primary studies that have evaluated return to practice schemes; (2) review of reviews of factors that influence retention; (3) **review of reviews of interventions for supporting recruitment and retention**. A decision was made that option 3 would be useful to inform practice.

## 1. BACKGROUND

This Rapid Evidence Map is being conducted as part of the Wales COVID-19 Evidence Centre Work Programme. The above question was suggested by the Royal College of Surgeons, Edinburgh.

### 1.1. Purpose of this review

National Health Service (NHS) waiting times have significantly increased over the past couple of years, particularly since the emergence of COVID-19, as elective and non-emergency treatments have been suspended or delayed to focus on the pandemic response. As of September 2021, 5.8 million people were waiting for their treatments to start in England, out of which 300,000 people have been on a waiting list for over a year, and 12,000 for over two years (Nuffield Trust 2021). In addition, emergency department waiting times reached a record high, with one in four people waiting longer than four hours for a decision on admission or discharge from the hospital (Nuffield Trust 2021). In Wales, treatment waiting times follow a similar tendency, with 240,306 people waiting more than 36 weeks for treatment from referral (Welsh Government 2021). Regarding emergency department visits in Wales, people were waiting a median of 3 hours and 7 minutes for admission to or discharge from hospital in October 2021. This waiting was an increase from 3 hours and 2 minutes in September 2021 (Welsh Government 2021).

One of the main reasons behind increasing waiting times and clearing the backlog is the NHS workforce shortage in every speciality, with 93,000 job vacancies UK-wide (Health and Social Care Committee 2021). Workforce shortages are worsened by the observable tendency, dated prior to the pandemic, that lower number of healthcare professionals enter the NHS than the number of qualified workers leaving (Health Committee 2018). Several factors can contribute to NHS staff retention issues, which can vary between different professional groups, for example workload pressures, poor access to continuing professional development, not feeling valued, and pay restraints (Health Committee 2018). Moreover, the pandemic response and Brexit put extra pressure on the NHS overstretching an already limited staff. Staff shortages not only affect the backlog, but impact on healthcare professionals’ health and wellbeing. Prior to the COVID-19 pandemic, one third of doctors reported feeling burnout. However, a survey conducted in England shows that following the start of the pandemic, 92% of the responding chairs and chief executives working in NHS Trusts reported concerns over their staff’s wellbeing, stress, and burnout (NHS Providers 2020).

Previous policy initiatives have focused on attempting to increase workforce numbers. For example, new routes to enter nursing were implemented, such as apprenticeships and fast track programmes, although these do not necessarily address shortages in the areas of mental health nursing and learning disabilities (Health Committee 2018). There is concern that existing recruitment and retention strategies are not sufficient enough to fill the workforce gap (Health and Social Care Committee 2021). Given that overseas or training options take too long to be able to help in the immediate short term, approaches to get clinicians and other health professionals who have left the medical profession to return gaps are needed. The Bringing Back Staff (BBS) programme (NHS 2021a) which started in March 2020, in response to the COVID-19 pandemic saw over 4000 clinicians returned to employment to provide valuable support to health and social care in frontline acute services, continuing health care programmes, clinical trials, COVID-19 vaccine centres and other settings. The BBS and other return to practice schemes have been particularly important since March 2020, as they have formed a significant part of the COVID-19 pandemic response. However, due to skill fade, professionals often require training to return to practice. Return to practice schemes provide a valuable route to practice with 50% of healthcare professionals who re-entered the NHS to help the pandemic response, have expressed interest in staying in some capacity on a longer term (NHS 2020). Recruiting new staff, retaining the existing NHS clinical workforce, and making return to clinical practice more attractive will require a number of approaches (BMA 2021). In order to inform practice, a rapid review of the best available evidence of innovations to attract, recruit, or retain NHS clinical staff is required. However, the extent of the available evidence is unclear, and may be limited to unpublished or grey literature. The aim of this rapid evidence map is to describe the extent and nature of the available evidence base for innovations (including return to practice) that could help attract, recruit, or retain NHS clinical staff, in order to identify the priorities and actions for a rapid review.

## 2. RESEARCH QUESTION

The eligibility criteria for the rapid evidence map, based on the Population, Phenomenon of Interest, Context, Study design (PiCoS) framework, are presented in Table 1.

**Table 1:**
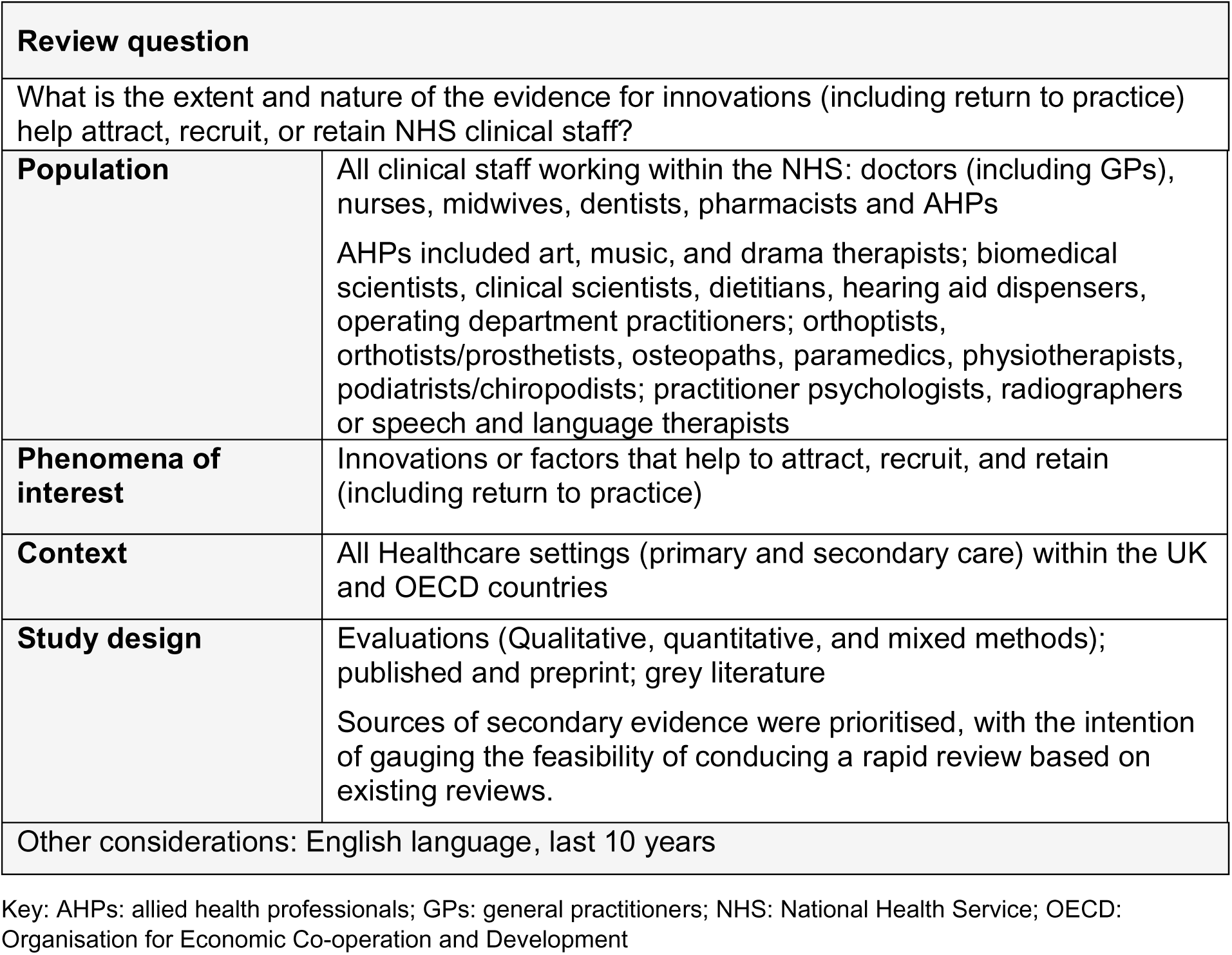
Eligibility criteria for the rapid evidence map.

## 3. SUMMARY OF THE EVIDENCE BASE

### 3.1. Type and amount of evidence available

A summary of the extent and type of evidence identified for different clinical staff working within the NHS, including nurses and midwives, doctors (including GPs), dentists, mixed groups of health professionals and allied health professionals (AHPs), is presented in Table 2.

**Table 2:** Summary of the type and amount of evidence.

| Evidence type | Total identified | Clinical staff working within the NHS |
| --- | --- | --- |
| Systematic reviews (SRs) | 13<br>11<br>1<br>3<br>5 | Nurses and midwives<br>Doctors (inc. GPs)<br>Dentists<br>AHPs<br>Mixed groups of health professionals |
| Rapid reviews (RRs) | 0<br>1<br>0<br>0<br>1 | Nurses and midwives<br>Doctors<br>Dentists<br>AHPs<br>Mixed groups of health professionals |
| Protocols<br>For work that is underway | 0<br>0<br>0<br>0<br>0 | Nurses and midwives<br>Doctors<br>Dentists<br>AHPs<br>Mixed groups of health professionals |
| Narrative reviews (NRs) | 4<br>3<br>0<br>1<br>3 | Nurses and midwives<br>Doctors<br>Dentists<br>AHPs<br>Mixed groups of health professionals |
| Rapid scoping reviews<br>(Rapid ScRs) | 0<br>1<br>0<br>0<br>0 | Nurses and midwives<br>Doctors<br>Dentists<br>AHPs<br>Mixed groups of health professionals |
| Scoping reviews (ScRs) | 2<br>2<br>0<br>1<br>1 | Nurses and midwives<br>Doctors<br>Dentists<br>AHPs<br>Mixed groups of health professionals |
| Umbrella reviews (URs) | 2<br>1<br>0<br>0<br>2 | Nurses and midwives<br>Doctors<br>Dentists<br>AHPs<br>Mixed groups of health professionals |
| Primary Studies | 5<br>7<br>0<br>5<br>1 | Nurses and midwives<br>Doctors (inc. trainees, GPs)<br>Dentists<br>AHPs (inc. pharmacist, occupational therapists)<br>Mixed groups of health professionals |
| Organisational reports | 1<br>0 | Nurses and midwives<br>Doctors |
|  | 0<br>0<br>1 | Dentists<br>AHPs<br>Mixed groups of health professionals |
| Organisational websites | 0<br>2<br>0<br>1<br>0 | Nurses and midwives<br>Doctors<br>Dentists<br>AHPs<br>Mixed groups of health professionals |

Key: AHPs: allied health professionals; GPs: general practitioners; NHS: National Health Service

Below the description of the evidence is presented by the phenomena of interest: return to practice, factors influencing recruitment and/or retention and interventions or strategies for improving recruitment and retention for each of the different professional groups.

#### 3.1.1. Return to practice

##### Nurses and midwives

• **One scoping review** investigated return to practice for nurses following a leave of absence for mental health reasons.
• **One systematic review** looked at return to practice interventions or bridging programmes for overseas trained nurses who would like to practice in Australia, Canada, New Zealand, or the UK.
• **One organisation report** explored **return to practice courses** for nurses.
• **Four primary studies** two of which were conducted in the UK (none of which were part of the scoping or systematic review) were identified for nurses returning to practice.

##### Doctors (including general practitioners)

• **Two systematic reviews** and **one narrative review** explored issues around doctors returning to practice.
• **Two organisational websites** (https://www.healthcareers.nhs.uk/explore-roles/doctors/returning-medicine and https://www.england.nhs.uk/coronavirus/returning-clinicians/ describe a range of initiatives that are available to help **clinicians** and **trainees,** who have been out of medical practice for a period of time, regain their skills and return to the NHS.
• **Four primary studies** conducted in the UK (none of which that were part of the systematic or narrative reviews) were identified for doctors returning to practice and an **additional three studies** for trainee doctors returning to their course of study.

##### Dentists

• **No systematic reviews** or **primary research evidence** were found for **return to practice** for **dentists.**

##### Allied health professionals

• **Two primary studies** investigated return to practice for **AHPs** across a range of specialities. One was conducted in the UK and evaluated a return to practice programme for AHPs and healthcare scientists. The other investigated flexible working arrangements for AHPs in Australia, returning to work after a period of maternity leave.
• **Two primary studies**, one conducted in New Zealand and one in Australia, were identified for **occupational therapists** returning to practice after a career break or after maternity leave.
• **One primary study** conducted in the UK was identified for **pharmacists** returning to practice after career breaks or moving sectors.
• **One organisational website** described (https://www.rpharms.com/) initiatives to support **pharmacists** returning to practice.

##### Mixed groups of healthcare professionals

• **One systematic review** looked at **return to practice** for a wide range of healthcare professionals.
• **One report** explored supporting staff to return to the NHS and return to practice schemes (NHS England 2020).
• **One primary study** in the UK (which was not part of the systematic review) was identified for mixed groups of health care professionals returning to practice.

#### 3.1.2. Factors influencing attraction, recruitment and retention

##### Nurses and midwives

• Eleven systematic reviews, two narrative reviews **and** one scoping review focused on factors affecting recruitment and/or retention of nurses.
• **One systematic review** was found for factors affecting recruitment and/or retention for **midwives.**

##### Doctors (including general practitioners)

• **One rapid review, two systematic reviews,** and **one scoping review** investigated factors influencing recruitment and/or retention of **doctors across a range of specialities.**
• **One umbrella review, three systematic reviews**, **one narrative review**, and **one rapid scoping review** investigated factors influencing recruitment and/or retention of **GPs.**

##### Dentists

• **No systematic reviews** were found that investigated factors influencing attraction, recruitment and retention for **dentists.**

##### Allied health professionals

• **Three systematic reviews** and **one narrative review** investigated factors that can influence recruitment and retention of **AHPs.**
• One of the systematic reviews specifically focused on **pharmacists,** and the other one on **physiotherapists and occupational therapists**. The narrative review focussed on **physiotherapists.**

##### Mixed groups of healthcare professionals

• **One umbrella review, one rapid review** and **three systematic reviews** investigated factors that influence retention of a wide range of professionals, including doctors, nurses, and AHPs.

#### 3.1.3. Interventions for improving attraction, recruitment and retention

##### Nurses and midwives

• **Two umbrella reviews**, **two narrative reviews,** and **one scoping review** explored interventions and strategies that could help retain existing nursing staff and recruit new professionals.
• **No systematic reviews** were found for **interventions for improving attraction, recruitment, and retention** for **midwives.**

##### Doctors (including general practitioners)

• **Two systematic review, one narrative review**, and **two scoping reviews** explored strategies that can help attract, recruit, and/or retain **physicians working in a wide range of disciplines.**
• **Two systematic reviews** (across three reports) and **one rapid scoping review** investigated strategies to help recruit and/or retain **GPs**.

##### Dentists

• **One systematic review** was found which explored **rural-exposure strategies** on the intention of dental students and dental graduates to practice in rural areas.

##### Allied health professionals

• **One narrative review** and **one scoping review** explored strategies that can help recruit, and/or retain **AHPs.** The narrative review investigated interventions for physiotherapists specifically, while the scoping review explored strategies for pharmacists.

##### Mixed groups of healthcare professionals

• **One umbrella review, one systematic review, one rapid review, one scoping review,** and **one narrative review** explored interventions and strategies that could help recruit and/or retain healthcare professionals.
• **Two narrative reviews** evaluated values-based recruitment.

### 3.2. Key findings

#### 3.2.1 Return to practice

There was limited secondary evidence available for return to practice, and further searches for primary studies were conducted to inform options for conducting a rapid review in this area. The included research of innovations for return to practice are reported in more detail in a series of tables (see Appendices 2 to 6), along with a summary of their key findings. An overview of the key findings are also provided here as a narrative.

##### Nurses and midwives

###### Evidence from systematic reviews

• **Academic bridging programmes** for internationally educated nurses which aim to help them **return to nursing** in the UK or other host countries, can provide experience and knowledge needed for their future work. However, some nurses considered these programmes as having no value, indicating that a fair and rational approach was needed when developing bridging programmes (Cruz et al. 2020).

###### Evidence from scoping reviews

• **Alternative to discipline programmes** (ADPs) are a humane approach to help nurses **return to practice** who have had a **leave of absence due to mental health issues**, mainly substance abuse (Covell et al. 2020). However, more research is needed on the effectiveness of interventions aiming to support nurses to return to work (Covell et al. 2020).

###### Evidence from organisational reports

• One organisation report identified the challenges in accessing return to practice (RTP) courses and made recommendations for future opportunities for RTP courses (Health Education England 2014b).

###### Evidence from primary studies

• The primary studies explored issues related to clinical training for military nurses (Kenward et al 2017), contact experiences and needs of nurses (Noorland et al 2021); views of primary care nurses (Ipsos Mori 2016) and satisfaction (McMurtrie et al 2014) of return to practice schemes.
• One study which was conducted in the Netherlands took place during the COVID-19 pandemic (Noorland et al 2021).

##### Doctors (including general practitioners)

Information from organisational websites provide links to further information for the Career Refresh for Medicine (CaReForMe), Supported Return to Training (SuppoRTT), Less Than Full Time Training (LTFT), medical support workers and the GP International Induction and Return to Practice Programmes.

• **CaReForMe** aims to help **doctors** who left the profession or took a career break, return to their practice easily and safely via different courses, e-learning, and additional supernumerary time (HEE 2021).
• **SuppoRTT** aims to support **trainee doctors** who are taking approved time out to return and complete their training. SuppoRTT offers enhanced supervision, mentoring, refresher courses, and online resources among other interventions (HEE 2020).
• **LTFT** is a flexible approach offering part-time training for **eligible trainee doctors and dentists** for numerous reasons, including disability, caring responsibilities, and personal or professional development opportunities (NHS 2021b).
• **Medical support workers** are clinicians who have acquired medical qualification but require supervision due to being out of practice for over a year or awaiting General Medical Council registration (NHS 2021c).
• The **GP Return to Practice programme 2021** offers multiple pathways to fit the needs of GPs wishing to return, such as taking a Learning Needs Assessment, submission of a portfolio, Medical Performers List refresher, and the Emergency Registered Practitioner Returner Programme (NHS 2021d).
• The **GP International Induction Programme 2021** provides a supported pathway for overseas qualified GPs to be inducted safely into NHS General Practice (NHS 2021d).

###### Evidence from systematic reviews

• There is limited evidence on how doctors’ skills are affected by time out of practice (GMC 2014).
• Skills might fade differently for different professionals in different settings (GMC 2014). However, evidence for skill fade is based on studies investigating the retention of various skills following training, rather than from research comparing skills before and after a career break (GMC 2014).
• In the US, it was found that the majority of re-entry physicians did not pursue additional training prior to returning to the workforce, unless it was required by medical or state specialty boards (Guth et al. 2020). While regulators have recently started increasing re-entry requirements, such as training or fitness to practice tests, meeting these changing regulations is the clinicians’ responsibility (Guth et al. 2020).

###### Evidence from narrative reviews

• A number of return to practice training schemes exist in the UK run by different professional bodies and training requirements vary depending on the amount of time spent out of practice (AoMRC 2012). The degree to which these training opportunities are mandatory is unclear. The most common period of absence, following which return to practice training is required, is one to two years (AoMRC 2012).

###### Evidence from organisational reports

• The NHS People Plan 2020/21 has a small section on encouraging former staff to return to the NHS and advice on supporting return to practice (NHS England 2020).

###### Evidence from primary studies

• One primary study analysed 1 year and 2 year evaluation data for SuppoRTT (HEE 2020).
• One primary study evaluated the Springboard initiative for physicians returning to training and two primary studies lengths and patterns of full time and LTFT training for trainee anaesthetist’s (Randive et al 2015) and another evaluated the accessibility and experiences of flexible training for trainee surgeons (Harries et al 2016).
• One primary study conducted an evaluation of the GP Returner (Induction and Refresher scheme) with a focus on practice placements (Morrison 2012).

##### Allied health professionals

###### Evidence from organisational websites

Information from organisational websites provide links to further information on two initiatives that are available for **pharmacists** considering returning to practice: mentoring and work shadowing

• Mentoring is described as a relationship between mentor and mentee which facilitates pharmacists’ return to practice via sharing experiences and reflection. Mentoring can be short and long term, and the Royal Pharmaceutical Society advises the formalisation of a clear contract for development to monitor and review mentees’ progress. Pharmacists wanting to return to practice can register their need for mentoring on the Royal Pharmaceutical Society’s online mentoring database. (https://www.rpharms.com/resources/pharmacy-guides/returning-to-practice-guide/supporting-you-with-return-to-practice)
• Work shadowing is a temporary, unpaid, work-based experience that aims to help pharmacists with networking, building credibility, strengthening their CVs and personal statements, and increasing their confidence for interviews. Pharmacists aiming to return to practice via shadowing need to organise their own placements, via contacting potential employers, their previous connections or colleagues. (https://www.rpharms.com/resources/pharmacy-guides/returning-to-practice-guide/work-shadowing)

###### Evidence from primary studies

• One primary study conducted in New Zealand found that the conditions that enabled **occupational therapists** successful return to practice included a strong sense of professional connectedness (sense of belonging and social connectedness to the profession), professional identity, accessibility to resources, and flexibility of employment option (Dodds and Herkt 2013).
• One primary study conducted in Australia found that **occupational therapists** returning from maternity leave had to make compromises to achieve a work-life balance. However, feeling valued by management and colleagues helped occupational therapists feel comfortable and confident with the compromises made (Parcsi and Curtin 2013).
• One primary study conducted in Australia explored flexible working arrangements for **AHPs** following maternity leave. Based on a mixed-methods investigation AHPs returned to practice on a part-time basis following maternity leave, and they stayed part-time for an extended period of time (Hulcombe et al 2020).
• One primary study investigated **pharmacists’** experiences of returning to practice after a career break or moving from a different sector (Phipps et al 2013).

##### Mixed groups of healthcare professionals

###### Evidence from systematic reviews

• There are **some risks** associated with **healthcare professionals returning to practice**, which can occur at a staff, organisational, or regulator level. No risks were found at a service user level (Campbell et al. 2019).
• Factors negatively impacting on professionals’ intention to return to practice could be organisational, such as lack of placements, supervision, peer and employer support, and personal barriers, including breast feeding, age, gender, personal health, and marital status (Campbell et al. 2019).
• **Approaches** that could help professionals **return to practice** included refresher/induction/re-entry programmes, supervision, mentoring, clear policies and planning, and support aiming at the individual, such as social networking, and peer support (Campbell et al. 2019).

#### 3.2.2. Bottom line summary for return to practice

Return to practice was investigated after leaving for a variety of reasons such as mental and physical health issues, disability, maternity leave, caring responsibilities, personal or professional development opportunities, career break, moving sectors, and retirement. The evidence highlights the challenges surrounding healthcare professionals wishing to return to practice particularly with regard to “skills fade”. There are a number of routes for healthcare professionals to return to practice or training and these are well documented. However, data on their effectiveness are limited. With primary research and organisational reports from professional bodies mainly offering suggestions from a range of perspectives. There is a wealth of evidence from primary studies describe views and experiences of those returning to practice and factors influencing decisions to return to practice.

#### 3.2.3. Factors influencing attraction, recruitment and retention

An overview of the existing reviews that looked at factors influencing attraction, recruitment, and retention of different clinical staff is provided in Table 6. This also includes a brief summary of the focus of each review and the number of included primary studies. An overview of the key findings the systematic, rapid and umbrella reviews are also provided here as a narrative.

##### Nurses and midwives

###### Evidence from systematic reviews

• **Mental health nurses** encounter factors that are **unique** to working within the mental health field due to the nature of the work and the work environment (Adams et al 2021).
• **Transition to practice programmes** are important in the pathway to registration for enrolled nurses (Blay and Smith 2020).
• **Post-COVID-19-pandemic studies** focused more on **predicting nurses’ turnover intention** through the pandemic’s negative impact on their psychological wellbeing (Falatah 2021).
• Factors that influence **millennial generation nurses’** (born 1981 – 1996) intention to stay include strong leadership, advancement opportunities, alignment of organizational and personal values, good co-worker relationships, healthy work-life balance, recognition and cutting-edge technology (Keith et al 2021).
• Having good working relationships, being supported by management, forming relationships through a women’s pregnancy journey, enjoying and being passionate about their role are some of the factors that influence **midwives’ intention to stay** in the profession (Bloxsom et al 2019).
• Multiple interrelated dimensions reflecting personal, professional and place factors influence **nurses’ decision making to work in rural and remote settings** (MacKay et al 2021).
• large number of **individual (personal), role and organisational factors** have been reported to influencing the retention and/or a **nurses’** intention to stay and these include

`o` **Individual (personal):** family reasons (Al Zamel et al 2020), personal and demographic influences (Marufu et al 2021).
`o` **Role:** education and career advancement (Marufu et al 2021), job complexity (Nei et al 2015), job control (Nei et al 2015), job satisfaction (Brown et al 2013; Al Zamel et al 2020), work pressure or job strain (Chamanga et al 2020, Nei et al 2015), role tension (Nei et al 2015), lack of time to complete tasks leading to work/life imbalance (Brown et al 2013, Nei et al 2015), traumatic/stressful workplace experiences (Khan et al 2019), professional issues (Marufu et al 2021), support at work (Marufu et al 2021).
`o` **Organisational**: organisational culture and values (Brown et al 2013), organisational commitment (Brown et al 2013; Al Zamel et al 2020; Nei et al 2015), quality of the work environment (Al Zamel et al 2020, Khan et al 2019, Marufu et al 2021), bullying at work (Al Zamel et al 2020), feelings of being valued (Brown et al 2013, Chamanga et al 2020), reward/recognition (Nei et al 2015), financial remuneration (Marufu et al 2021), working conditions (Chamanga et al 2020), staffing levels (Marufu et al 2021), job security (Al Zamel et al 2020), supportive and communicative leadership (Nei et al 2015, Marufu et al 2021, Al Zamel et al 2020).
`o` Some of these factors are connected, interrelated and interchangeable within the main categories of individual, role, and organisational issues.

##### Doctors (including general practitioners)

###### Evidence from systematic reviews

• Factors governing rural recruitment and retention strategies for **doctors in rural areas within high income countries** include having a rural background, rurally focused education and training, personal and professional circumstances, and integration with the community, family-unit considerations for partners and children. Barriers to recruitment include concerns over isolation and poor perception of rural practice (Holloway et al 2020).
• Working conditions and financial factors are associated with the retention and willingness of **physicians to serve in rural and underdeveloped areas**. Recruiting physicians, who are from rural backgrounds and rural origins, is another determining factor in physicians’ retention (Mohammadiaghdam et al 2020).
• Long-term **recruitment and retention** of **doctors in remote areas of Australia and Canada** is influenced by a broad range of negative and positive perceptions and experiences (Koebisch et al 2020, Viscomi et al 2013, Wieland et al 2021). Some of the key factors identified are:

`o` Professional (including training), organisational and personal (Wieland et al 2021).
`o` Rural background (of medical student or partner, or both), male gender, interest in living in a rural area and meaningful rural elective exposure during medical training (Viscomi et al 2013).
`o` Scope of practice was deemed very important as a factor of recruitment, as was attraction to rural lifestyle (Koebisch et al 2020).
`o` Incentives were found to be of little importance (Koebisch et al 2020).
• **GPs** with rural backgrounds or rural experience during undergraduate or postgraduate medical training are more likely to practise in rural areas (Ogden et al 2020).

###### Evidence from umbrella reviews

• One hundred and fifty-eight factors influencing the recruitment and retention of **family physicians in rural areas** were identified and summarised into 11 categories. The three categories referenced most often were related to training, personal and practice which resemble three distinct phases of a family physician’s life: pre-medical school, medical school, and post-medical school (Asghari et al 2020).

###### Evidence from rapid reviews

• Factors influencing the retention of **doctors working in primary and secondary care** were identified as low morale, disconnect, unmanageable change, lack of personal and professional support, and feelings of mastery and membership (Andah et al 2021).
• A variety of individual-level, role-related, organisational / team-related and system-level factors affected retention of **anaesthetists** (RCOA 2021b).

##### Allied health professionals

###### Evidence from systematic reviews

• A large number of **organisational/workplace structure and personal factors** have been reported to influence the recruitment and/or retention of **AHPs working in Metropolitan, rural, and remote locations**. Career opportunities positively impacted on recruitment, while lack of opportunity negatively affected retention. Previous location exposure positively impacted recruitment however had limited impact on retention. Similarly, a diverse clinical load was reported as being attractive during recruitment, but unmanageable caseloads affected retention (Couch et al 2021).
• Factors associated with recruitment and retention of **pharmacists in rural practice** have been identified as geographic and family-related, economic and resources, scope of practice or skills development, the practice environment, and community and practice support factors (Terry et al 2021).
• Factors influencing recruitment and/or retention of **occupational therapists and physiotherapists**, and their decisions to locate, stay or leave rural communities was influenced by: availability of and access to practice supports, opportunities for professional growth and understanding the context of rural practice, such as larger caseloads, limited referral options, decreased access to resources and limited access to continuing education (Roots and Liu 2013).

##### Mixed groups of healthcare professionals

###### Evidence from systematic reviews

• A broad range of factors are associated with rural retention of **Australian primary healthcare workers,** and it was suggested that retention strategies should be multifaceted and bundled (Russell et al 2017).
• Factors influencing the retention or non-retention of **healthcare professionals** during or after a disaster are multifaceted and a combination of several appropriate strategies should be used to respond to this (Jamebozorgi et al 2021).

###### Evidence from umbrella reviews

• The main factors impacting retention for **healthcare workers in rural and remote areas** in developed and developing countries were opportunities for professional advancement, professional support networks and financial incentives. The most important factors influencing recruitment were rural background and rural origin, followed by career development (Mbemba et al 2016).

###### Evidence from rapid reviews

• There is a paucity of good evidence about the best ways to retain **professionals in the NHS**, strategies identified were peer support, reduced hours, bonuses and portfolio roles, however it is difficult to say whether these are effective (RCOA 2021b).

#### 3.2.4. Bottom line summary for factors

Factors influencing attraction, recruitment and retention of nurses, midwives, doctors, and AHPs can be organised into three main groups: individual (personal), role-related and organisational. Individual or personal factors can include family reasons and demographic characteristics. Role-related factors can encompass issues related to specific professions, support at work, job satisfaction, work pressures, work/life balance, and career progression among others. Organisational factors can comprise organisational culture, work environment, valuing staff, financial renumeration, staffing levels and supportive leadership among others. In relation to recruitment and retention to rural and remote areas, rural background/origin and rural training are factors often cited to influence healthcare professionals’ decisions.

#### 3.2.5. Improving attraction, recruitment and retention

An overview of the existing reviews that looked at interventions/strategies for improving attraction, recruitment, and retention of different clinical staff is provided in Table 7. This also includes a brief summary of the focus of each review and the number of included primary studies and the key findings. An overview of the key findings is also provided here as a narrative.

##### Nurses and midwives

###### Evidence from umbrella reviews

• One umbrella review, that investigated interventions to **reduce turnover** in **adult nursing** in hospital and community settings (mainly from the USA), reported evidence of the effect of a small number of interventions which decrease turnover or increase retention of nurses, these being **preceptorship of new graduates** and **leadership for group cohesion** (Halter et al. 2017).
• One umbrella review identified four broad types of interventions as potential strategies that could influence the retention of nurses in rural and remote areas: **education and continuous professional development** interventions, **regulatory** interventions, **financial** incentives, and **personal and professional support** (Mbema et al 2013).

###### Evidence from scoping reviews

• A scoping review showed that barriers to and strategies for millennial **nurse retention** commonly focus on **the work environment** and the **relationships between nursing leadership and the bedside nurse** (McClain et al 2022).

###### Evidence from narrative reviews

• Two narrative reviews examined the impact of nursing practice environment and found a wealth of evidence supporting the impact of a **positive practice environment** to support **nurse retention** (Redknap et al 2015; Twigg and McCullough 2014).

##### Doctors (including general practitioners)

###### Evidence from systematic reviews

• Two systematic reviews explored the effectiveness of interventions to improve recruitment and/or retention of doctors in **rural areas** and found that

`o` Successful strategies included student **selection from rural backgrounds** into medical school (Johnson et al 2018, Kumar and Clancy 2020) and undergraduate education programs and early postgraduate **training in a rural environment** (Kumar and Clancy 2000).
`o` **Bundled or multifaceted interventions** may be more effective than single factor interventions (Kumar and Clancy 2020).
`o` Other key potential rural predictors included: rural interest/intentions prior to the program, generalist practice intentions, an interest in primary care and family medicine, financial and rural bonded scholarships and the type and quality of a rural immersion experience and its duration (Johnson et al 2018).
• A systematic review investigating recruitment strategies for **GPs** in OECD countries noted that **studies are scarce**, with most focusing on remote rural locations (Peckham et al 2016, Marchand and Peckham 2017).
• One internationally focused systematic review found limited evidence for **GP retention initiatives** that focused on wellbeing, peer support, or support for professional development or research. Mixed evidence was found for **financial rewards** (Verma et al 2016).

###### Evidence from scoping reviews

• Two scoping reviews found a paucity of evidence that directly addressed efforts to improve **retention of doctors** with **studies mainly offering suggestions** from a range of perspectives.

`o` For doctors working in **emergency medicine** suggestions included improving workflow and staffing, self-care and compassion dialogues, and work scheduling (rostering) (Darbyshire et al 2021).
`o` For **paediatricians,** the most important strategies employed to enhance recruitment and retention include professional advocacy, workforce diversity, mentorship, improving working conditions, career flexibility and enhancing educational opportunities (Mallett et al 2021).
• One rapid scoping review offered suggestions that could improve **recruitment and retention of GPs**. For **recruitment**, suggested strategies included improved funding for clinical placements, encouragement of respect between medical professionals, inspiring GP role models and leaders, improving the public image of general practice through outreach work in schools and with the public. For **retention**, strategies related to trying to increase capacity and reduce workload, encourage variation in working life through portfolio careers and sub-specialisms as well as greater support for those wishing to change their clinical workload (Mitchell et al 2018).

###### Evidence from narrative reviews

• A narrative review that focused on rural areas (preventative as opposed to curative services) suggested that **continuing medical education** activities show promise as a strategy to recruit and retain physicians in **less attractive specialties** (Thi Nguyen et al 2021).

##### Dentists

###### Evidence from systematic reviews

• One systematic review of **rural-exposure strategies** for dental students and graduates found that enrolling students with **rural backgrounds** and imposing **compulsory clinical rotation in rural areas** during their study appeared to be effective in tackling the shortage and maldistribution of dentists in rural areas (Suphanchaimat et al 2016).

##### Allied health professionals

###### Evidence from scoping reviews

• One scoping review focused on **pharmacists in rural and remote** Australia suggested strategies to increase the workforce and included: enrolment of **students from rural backgrounds**, availability of **support personnel** for rural initiatives, **extended rural placement** and the inclusion of **rural content in the teaching** curriculum (Obamiro et al 2012).

###### Evidence from narrative reviews

• A narrative review in Australia suggested strategies to improve retention of skilled **physiotherapists** which were broadly grouped into improving **professional support** in the workforce and **assisting the re-entry process** for physiotherapists seeking to return to the workforce (Pretorius et al 2016).

##### Mixed groups of healthcare professionals

###### Evidence from systematic reviews

• Only one study was included in the systematic review by Grobler et al (2015) which suggested that the implementation of a **National Health Insurance scheme** in Taiwan made medical care more affordable possibly leading to better geographical distribution of health care professionals.

###### Evidence from umbrella reviews

• One umbrella review focused on **rural health workers** and found that recruiting **rural students and rural placements** improved attraction and retention, although most studies were without control groups, which made conclusions on effectiveness difficult (Esu et al 2021).

###### Evidence from rapid reviews

• A rapid review of strategies for improving retention for a range of health professionals identified few studies on **anaesthetists or surgeons** with those available focusing on improving **mental wellbeing or job satisfaction**. For **other health professionals**, interventions which fed into the following categories were identified: support initiatives, professional development, reimbursement and terms, other initiatives (RCOA 2021b).

###### Evidence from scoping reviews

• One scoping review focused on strengthening human resources (healthcare workers) in epidemics recommended that decision makers should implement strategies that cover five themes (**preparation, protection, support, care, and feedback**) which are adjusted to context. In addition to the main themes, fifteen sub themes were also identified (Jelyani et al 2021).

###### Evidence from narrative reviews

• One narrative review evaluated **value-based recruitment** in the UK NHS and argued that insights regarding the impact of value congruence between employees and organisations should be interpreted with caution, as outcomes may not be immediately generalisable to a healthcare context and in particular to the NHS, due to different organisational drivers in other organisations which are focused on job satisfaction, productivity and reduced staff turnover as opposed to providing best possible patient care (HEE 2014a).
• Theoretical implications of one narrative review focused on **recruiting for values** in healthcare and healthcare education suggested that prosocial implicit trait policies, which could be measured by selection tools such as situational judgement tests and multiple mini interviews, may be linked to individuals’ values via effective behaviours considered in given situations (Patterson et al 2016).
• A narrative review covering 20 European countries including the UK found a lack of evidence about whether strategies to recruit were effective and suggested single recruitment interventions on their own have little impact**, bundles of interventions** are more effective (Kroezen et al 2015).

#### 3.2.6. Bottom line summary for effectiveness

A number of reviews have investigated interventions/strategies for improving attraction, recruitment, and retention of different clinical staff, many of which focused on rural and remote areas.

### 3.3. Areas of uncertainty

Remaining uncertainties include:

• There were no systematic reviews that evaluated the CaReForMe, SuppoRTT, LTFT medical support workers and the GP International Induction and Return to Practice Programmes.
• There were no systematic reviews for AHPs that evaluated return to practice schemes.
• With the exception of occupational therapy and pharmacy there were no primary studies across the AHPs that evaluated return to practice schemes.
• Across all disciplines there are a number of websites, reports, guidance, other non-research publications that addresses healthcare staff returning to practice pre-pandemic after a career break, mental health issue or maternity leave. However, there is limited systematic review evidence.
• Across all disciplines there is very little systematic review evidence in relation to staff returning to practice post retirement as a result of the COVID-19 pandemic.
• A systematic review by Campbell et al (2019), although addressing return to practice across a number of healthcare professions, did not provide their search strategy and did not respond to an email request, and the findings were presented as a pooled narrative across all the professional groups included.
• Across all disciplines there are a number of websites, reports, guidance and other non-research publications that address the challenges and concerns relating to the workforce issues that have been present across both pre and post COVID-19 pandemic. However, the number of research studies evaluating strategies to improve recruitment, retention, and return to practice for healthcare professionals is low.
• From the reviews presented in this rapid evidence map, only three aimed to investigate attraction of healthcare professionals (Hutchinson et al 2012, Esu et al 2021, Viscomi et al 2013). All three reviews, out of which two focussed on rural and remote areas (Esu et al 2021, Viscomi et al 2013), found limited evidence on healthcare professionals’ attraction across all disciplines.

### 3.4. Options for further work

#### Option 1

A rapid review of primary studies that have evaluated return to practice schemes.

##### Eligibility criteria for Option 1

• **Participant groups** Doctors (including GPs and trainees), and nurses, as this rapid evidence map uncovered primary research investigating RTP schemes for these professionals. If stakeholders are interested, there is also potential for including occupational therapists, pharmacists, medical specialties, such as paediatrics, anaesthetics, and surgery, and research demonstrating pooled findings from RTP schemes for mixed groups of healthcare professions.
• **Interventions: Return to practice** schemes supporting healthcare professionals returning following career change or maternity leave. If stakeholders are interested, investigating RTP schemes focusing on support for professionals after a career break due to ill health (including mental health issues) is also possible. If stakeholders are interested, studies looking at factors influencing RTP, and research exploring healthcare professionals’ characteristics, views, and experiences with RTP schemes as part of an evaluation could also be included.
• **Comparisons:** All comparison groups would be included that are presented across the studies.
• Outcomes: **Potential outcomes could be r**ecruitment numbers, professionals’ skills, abilities, and performance following attendance in RTP schemes. Moreover, studies investigating risks associated with RTP would also be included.
• **Study designs:** Evaluations (quantitative, qualitative, or mixed methods)
• O**ther considerations:**

`o` English language publications published in the past 10 years with a particular focus on studies from the UK.
`o` Pre-prints, peer reviewed papers, and grey literature

##### Exclusion criteria

• The following participant groups would be excluded due to the lack of evidence found in the rapid evidence map: dentists, and AHPs (apart from occupational therapists, and pharmacists). Research focusing on student nurses and midwives would also be excluded, as their return to their studies require a different approach than qualified professionals RTP schemes.
• Interventions that focus on healthcare professionals supporting patients to return to work after an illness or on reopening of clinics and dental practices.
• Outcomes that focus on healthcare professionals’ intentions to retire.
• Study designs that only contain descriptions of interventions but not an evaluation.
• Guidelines of professional organisations

#### Option 2

A rapid review of reviews of factors that influence retention.

##### Eligibility criteria

• **Population:** Doctors (including GPs), nurses, midwives, dentists, pharmacists, physiotherapists, occupational therapists, and paramedics as this rapid evidence map uncovered secondary research on factors that influence retention of these healthcare professionals. If stakeholders are interested, there is also potential for including medical specialties, such as paediatrics, anaesthetics, and surgery, radiographers, and research demonstrating pooled findings for mixed groups of healthcare professions.
• **Phenomena of interest:** Factors that influence retention of healthcare professionals in their current place of work. If stakeholders are interested, investigating factors affecting retention in rural and remote areas is also possible. However, based on the rapid evidence map, there are a large volume of publications that identify the key factors affecting retention in rural and remote healthcare settings.
• **Context:** All healthcare settings.
• **Study designs:** Systematic reviews and rapid reviews. Preprints and published peer-reviewed papers
• **Other considerations:**

`o` English language publications published in the past 5 years.
`o` Pre-prints and peer reviewed papers

##### Exclusion criteria

• All other AHPs would be excluded due to the lack of evidence found in the rapid evidence map. Research focusing on factors influencing retention of students would also be excluded, as their retention is affected by different issues than those of qualified professionals.
• The following study designs would be excluded: narrative reviews, scoping reviews, and review protocols.

#### Option 3

A rapid review of reviews of interventions for supporting recruitment and retention

##### Eligibility criteria

• **Population:** Doctors (including GPs), nurses, midwives, dentists, pharmacists, physiotherapists, and occupational therapists as this rapid evidence map uncovered secondary research on interventions supporting recruitment and retention of these healthcare professionals. In addition, the following AHPs are included, as they are professions on the UK Visas and Immigration (2021) shortage occupation list: psychologists, paramedics, radiographers, radiotherapists, speech and language therapists.
• **Phenomena of interest: I**nterventions supporting recruitment and retention of healthcare professionals. In addition, interventions aiming at recruiting students into healthcare jobs would also be included. The included secondary research needs to focus on the evaluation of these interventions.
• **Context**: All healthcare settings, including rural areas, which might be of relevance to Wales.
• **Study designs**: Systematic reviews (summarising mixed methods, qualitative, and quantitative studies with robust evaluations), rapid reviews, and scoping reviews that include an evaluation component. Scoping reviews could be included as the rapid evidence map indicates that numerous scoping reviews are available.
• **Other considerations:**

`o` English language publications published in the past 5 years.
`o` Pre-prints and peer reviewed papers

##### Exclusion criteria

• All other AHPs not mentioned above would be excluded due to the lack of evidence found in the rapid evidence map. Research focusing on interventions influencing recruitment and retention of students would also be excluded, as their recruitment and retention for university courses requires a different approach.
• Transition programmes for newly qualified nurses, such as mentoring, preceptorship, and residency programmes, would be excluded.
• The following study designs would be excluded: narrative reviews and review protocols.

## 4. NEXT STEPS

The findings of this rapid evidence map were presented at a Stakeholder meeting (held on the 2nd February 2022) and the intended focus of the planed subsequent rapid review was discussed. A decision was made that the rapid review should focus on the following research question, and that this would be based on an overview of existing reviews (option 3):

RR00028. Rapid Review of what are the effectiveness of interventions/innovations relevant to the Welsh NHS context to support recruitment and retention of clinical staff. April 2022. This report can be accessed in the WCEC library: https://healthandcareresearchwales.org/about-research-community/wales-covid-19-evidence-centre

## 5. RAPID EVIDENCE MAP METHODS

### 5.1. Evidence sources

COVID-19 specific and general repositories of evidence reviews, websites of key third sector and government organisations and three databases (PubMed, Medline and Cinahl) were searched for English language publications for the last 10 years (conducted in January 2022). An audit trail of the search process is provided within the resource list (Table 3). Due to the rapid nature of this work not all third sector and government organisations were searched but a note was made of their existence for future reference.

**Table 3:**
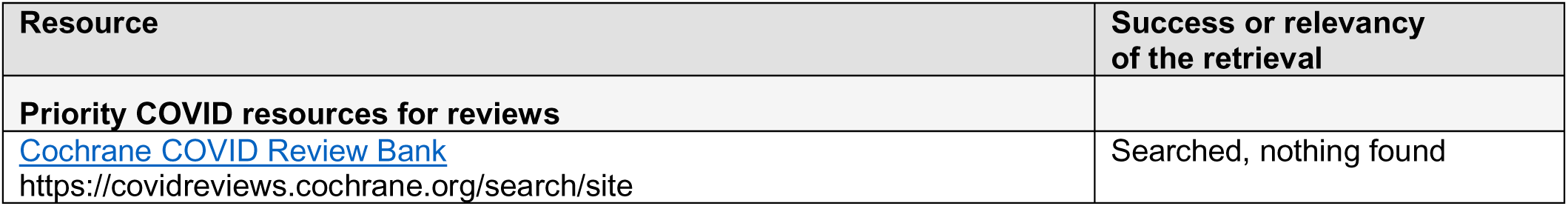
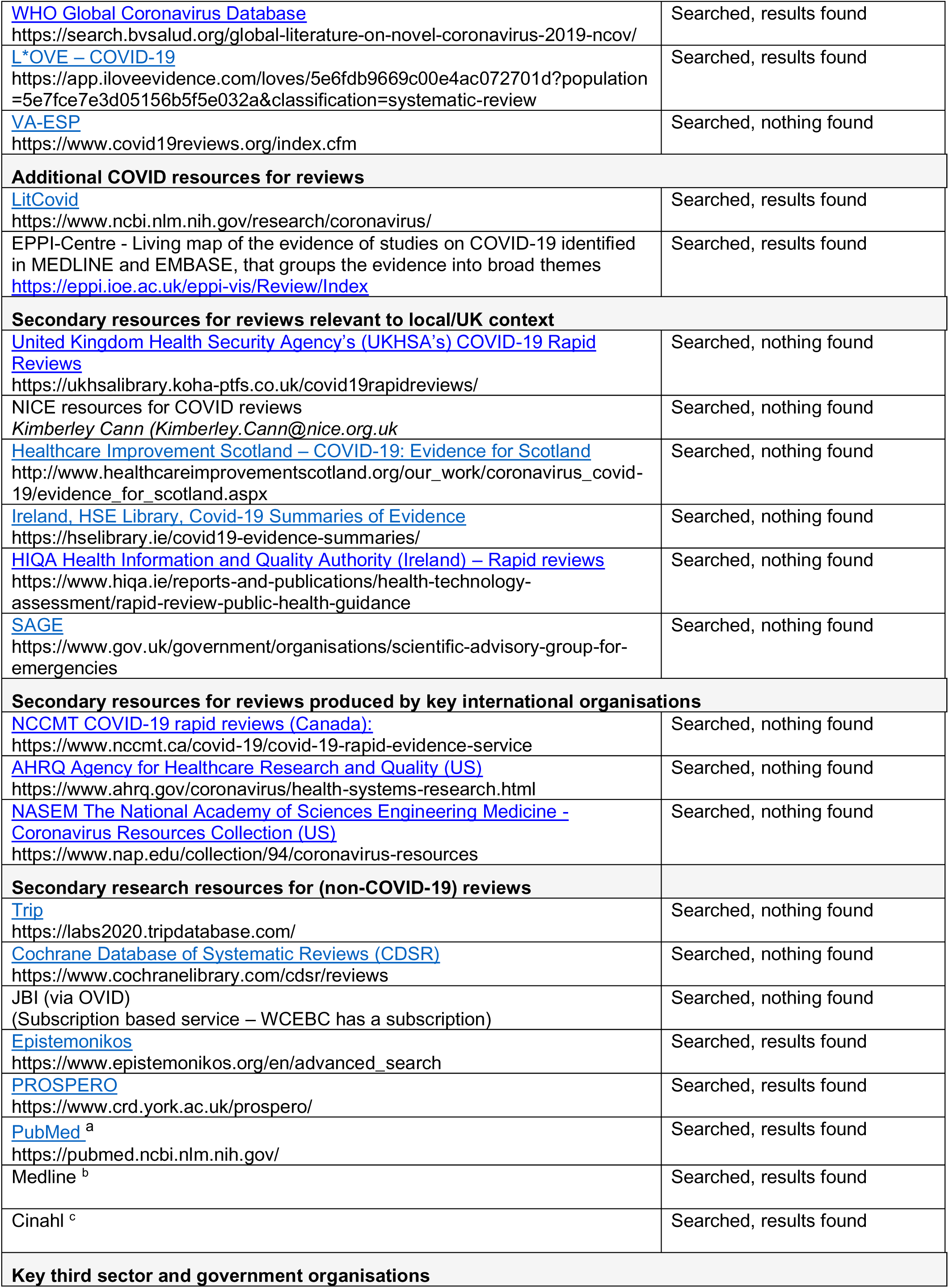
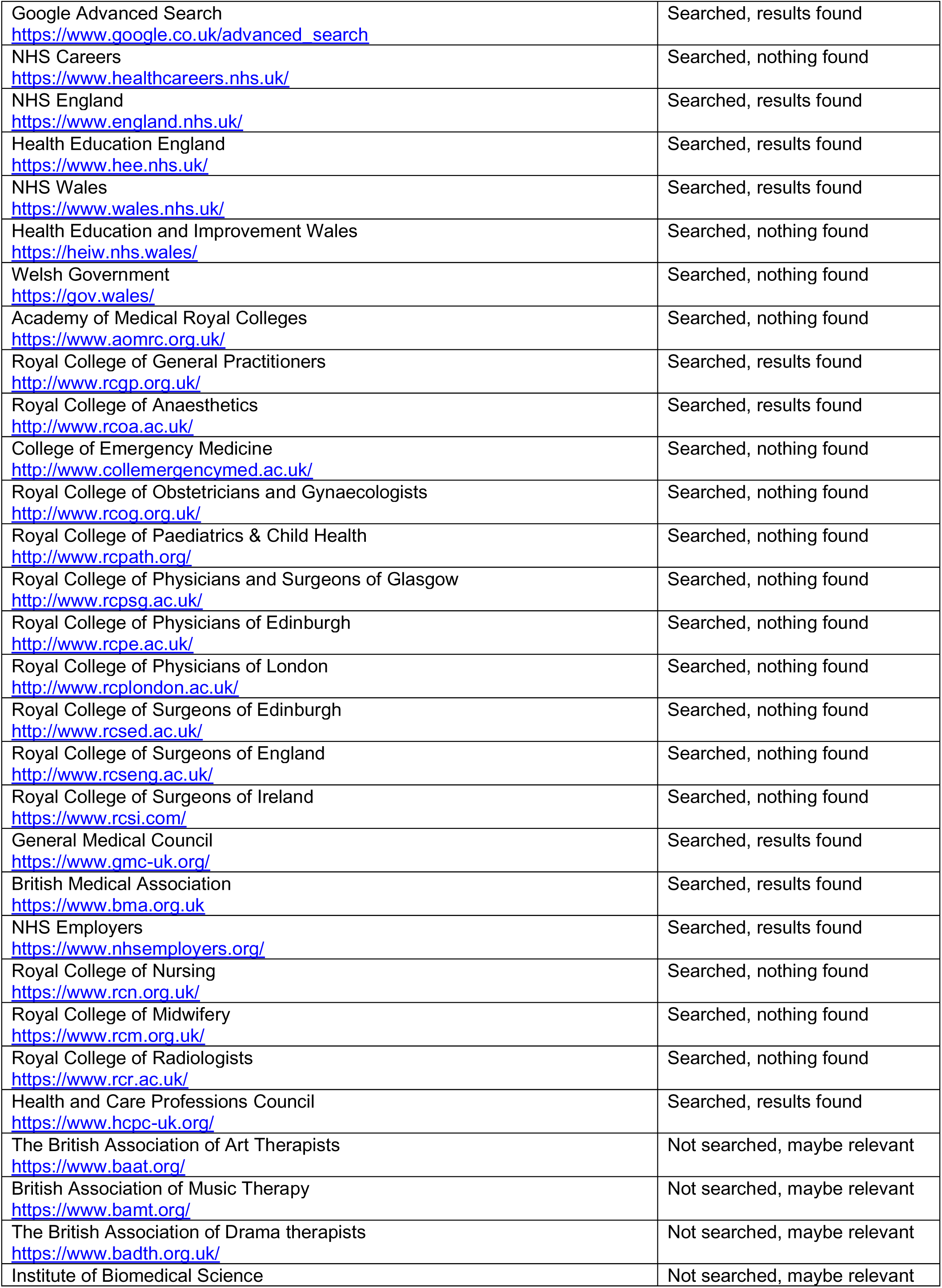
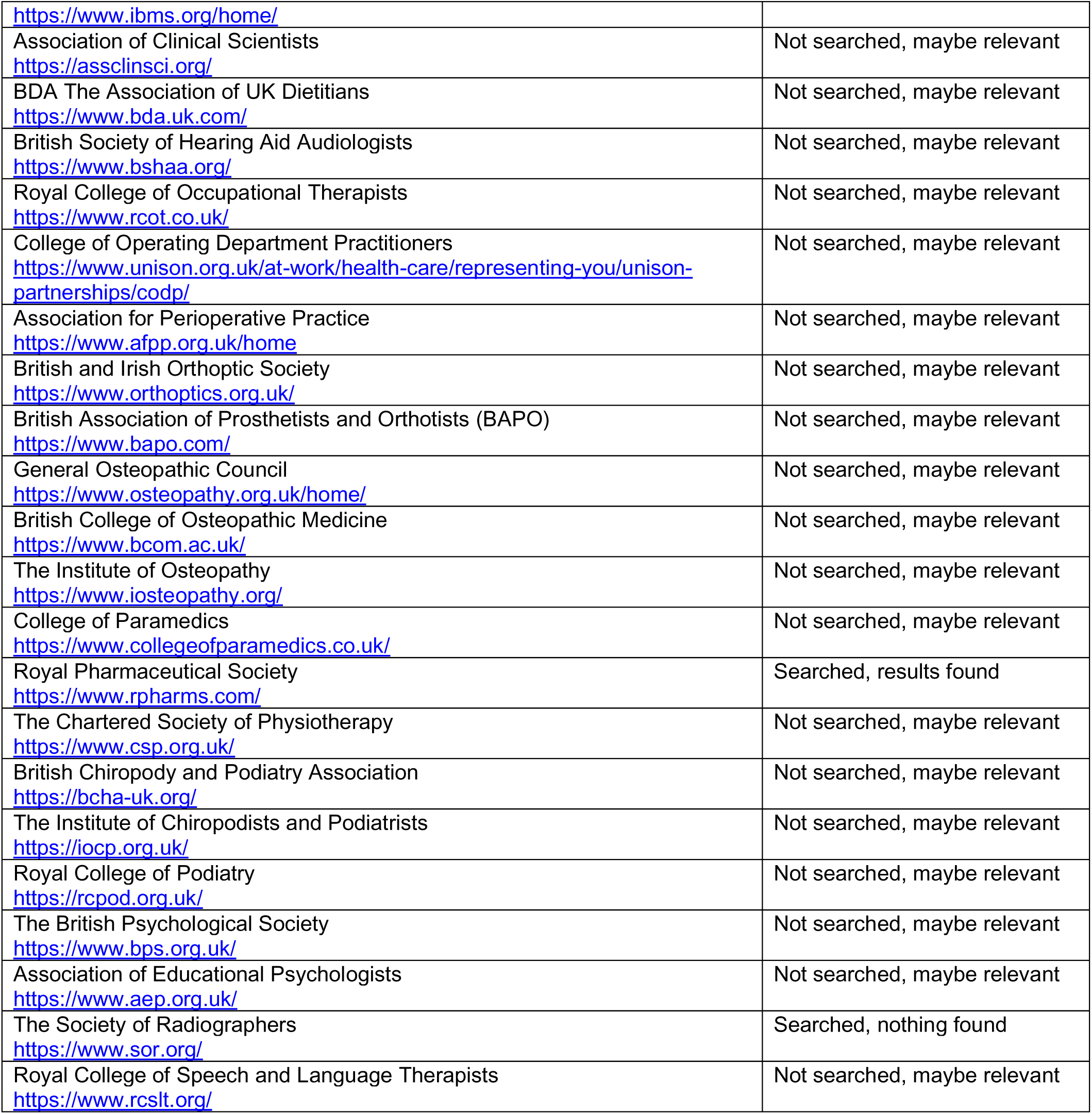
List of resources searched.

### 5.2. Search strategy

Searches were limited to English-language publications and did not include searches for primary studies if secondary research relevant to the question was found. The search strategies are listed in Appendix 1.

### 5.3. Reference management

All citations retrieved from the database searches were imported or entered manually into EndNote^TM^ (Thomson Reuters, CA, USA) and duplicates removed. Irrelevant citations were removed by searching for keywords within the title using the search feature within the Endnote software. At the end of this process the citations that remained were exported as an XML file and then imported to Covidence^TM^.

### 5.4. Study selection process

Search hits were screened for relevance by a single reviewer using the information provided in the title and abstract using the software package Covidence^TM^. For citations that appeared to meet the inclusion criteria, or in cases in which a definite decision could not be made based on the title and/or abstract alone, the full texts of all citations were retrieved. The full texts were screened for inclusion by two reviewers using the software package Covidence^TM^ and any disagreements resolved by a third reviewer. Priority was given to robust evidence synthesis using minimum standards (systematic search, study selection, quality assessment, appropriate synthesis). The reference lists of included umbrella reviews and rapid reviews that included systematic reviews were scanned for additional references.

### 5.5. Data extraction

For the evidence for return to practice data extraction was conducted by one reviewer and checked for accuracy by a second reviewer. For factors and interventions/strategies for improving attraction, recruitment and retention data extraction was conducted by one reviewer from the information contained within the abstract. Where information was missing or incomplete in the abstract, the full-text article was consulted.

### 5.6. Assessment of methodological quality

Formal quality appraisal of the included secondary evidence was not conducted.

### 5.7. Additional searches for primary studies

As secondary evidence was limited for return to work, a further targeted search for primary studies was conducted to inform options for further work. The same search terms were used as presented in Appendix 1 for Medline, Cinahl and PubMed but without restricting to reviews and additional searches for specific return to work programs (“Supported Return to Training Programme” or SuppoRTT, “Career Refresh for Medicine programme” or CaReForME, “Less than full time training”, “Bring back staff programme*”, Medical support worker*, “Return to practice programme*”). Findings from such studies have not been tabulated but an indication is given of the amount of literature for different aspects of the question.

### 5.8. Data summary

The data was presented in tables and summarised as a series of bullet points by type of evidence for each professional group.

## Data Availability

All data produced in the present work are contained in the manuscript

## Abbreviations

Acronym: Full Description

ADP: Alternative to discipline

AHP: Allied health professionals

AoMRC: Academy of Medical Royal Colleges

BBS: Bring Back Staff

BMA: British Medical Association

CaReForMe: Career Refresh for Medicine

CV: Curriculum Vitae

GMC: General Medical Council

GPs: General Practitioner

HEE: Health Education England

HEI: Higher Education Institution

IEN: Internationally educated nurses

LTFT: Less Than Full Time

NHS: National Health Service

NR: Narrative review

OECD: Organisation for Economic Co-operation and Development

PRISMA: Preferred Reporting Items for Systematic Reviews and Meta-Analyses

RCOA: Royal College of Anaesthetists

RR: Rapid review

RTP: Return to practice

ScR: Scoping review

SR: Systematic review

SuppoRTT: Supported Return to Training

UK: United Kingdom

UR: Umbrella review

US/USA: United States/United States of America

## 7.2. Acknowledgements

The authors would like to thank our stakeholders Luke Davies, Charlette Middlemiss, Ian Owen, Sally Anstey and members of the Wales COVID-19 Evidence Centre for their advice and guidance.

## 8. ABOUT THE WALES COVID-19 EVIDENCE CENTRE (WCEC)

The WCEC integrates with worldwide efforts to synthesise and mobilise knowledge from research.

We operate with a core team as part of Health and Care Research Wales, are hosted in the [Wales Centre for Primary and Emergency Care Research (PRIME)](http://www.primecenter.wales/), and are led by [Professor Adrian Edwards of Cardiff University](https://www.cardiff.ac.uk/people/view/123022-edwards-adrian).

The core team of the centre works closely with collaborating partners in [Health Technology Wales](https://www.healthtechnology.wales/), Wales Centre for Evidence-Based Care, [Specialist Unit for Review Evidence centre](https://www.cardiff.ac.uk/specialist-unit-for-review-evidence), [SAIL Databank](https://healthandcareresearchwales.org/about-research-community/secure-anonymised-information-linkage-sail-databank), [Bangor Institute for Health & Medical Research/Health and Care Economics Cymru](https://www.bangor.ac.uk/health-sciences/research/index.php.en), and the [Public Health Wales Observatory](https://phw.nhs.wales/services-and-teams/observatory/).

Together we aim to provide around 50 reviews per year, answering the priority questions for policy and practice in Wales as we meet the demands of the pandemic and its impacts.

### Director

Professor Adrian Edwards

### Contact Email

**Website:** https://healthandcareresearchwales.org/about-research-community/wales-covid-19-evidence-centre

All reports can be downloaded via the library on the WCEC website.

## 9. APPENDICES

## 9.1 Appendix 1: Search strategies

**^a^ Details of Pubmed searches (conducted 26^th^ January 2022)**

*Pubmed search for return to practice*

(return*[Title] OR re-ent*[Title] OR reent*[Title] OR re-licensure[Title] OR relicensure OR reactivat*[Title] OR revalid*[Title] AND

nurs*[Title] OR midwi*[Title] OR medic*[Title] OR practice[Title] OR practise[Title] OR NHS[Title] OR healthcare[Title] OR NHS[Title] OR doctor*[Title] OR clinician*[Title] OR physician*[Title] OR surgeon*[Title] OR dentist*[Title] OR allied health profesional[Title]

AND (review[Filter] OR systematicreview[Filter]) Filters: Review, Systematic Review, in the last 10 years, English

Nurses **(7 hits)**

Midwives **(23 hits)**

Doctors **(47 hits)**

General practitioners **(3 hits)**

Dentists **(0 hits)**

Allied health professionals **(0 hits)**

*Pubmed searches for attract, recruit and retain*

(retain*[Title] OR attract*[Title] OR recruit*[Title] OR retention*[Title] AND

nurs*[Title] OR midwi*[Title] OR medic*[Title] OR practice[Title] OR practise[Title] OR NHS[Title] OR healthcare[Title] OR NHS[Title] OR doctor*[Title] OR clinician*[Title] OR physician*[Title] OR surgeon*[Title] OR dentist*[Title] OR allied health professional[*Title]

AND (review[Filter] OR systematicreview[Filter]) Filters: Review, Systematic Review, in the last 10 years, English

Nurses **(49 hits)**

Midwives **(17 hits)**

Doctors **(39 hits)**

General practitioners **(11 hits)**

Dentists **(2 hits)**

Allied health professionals **(0 hits)**

**^b^ Details of Medline searches from 2012 to current and limited to reviews (conducted 24^th^, 31^st^ January 2022)**

*Medline searches within titles and abstract for attract, recruit and retain* (recruit* or retain* or retention or attract*) adj10

- (nurs*) **(242 hits)**
- (midwives or midwifery)). **(9 hits)**
- (doctor* or clinician* or physician* or surgeon* or healthcare or health care or NHS or national health service*) **(387 hits)**
- (clinical practice* or consultant*) **(16 hits)**
- (GP* or general practitioner* or general practice or primary care) **(143 hits)**
- (dentist* or dental*) **(14 hits)**
- (pharmacist* or pharmacy).**(30 hits)**
- “occupational therap*” **(8 hits)**
- (physiotherap* or “physical therap*”) **(34 hits)**
- (radiographer* or radiologist*) **(2 hits)**
- (“speech and language therap*”) **(0 hits)**
- (“practitioner psychologist*” or “registered psychologist*”) **(0 hits)**
- (chiropodist* or podiatrist*) **(0 hits)**
- (paramedic*) **(0 hits)**
- (osteopath*) **(0 hits)**
- (orthotist*) **(0 hits)**
- (orthoptist*) **(0 hits)**
- (“operating department practi* or ODP*) **(0 hits)**
- (dietician* or dietetics) **(0 hits)**
- (“clinical scientist*” or “biomedical scientist*”) **(0 hits)**
- (“art therap*” or “music therap*” or “drama therap*”) **(0 hits)**
- (AHP* or “allied health profession*” or “allied health workforce” or PAMs or “professions allied to medicine”) **(0 hits)**

*Medline searches within titles and abstracts for return to practice* ((return or re-ent* or reent* or re-licen* or relicen* or re-activate or reactivate or re-validate or revalidate or re-employ* or reemploy*) adj10

- (nurs*) **(14 hits)**
- (midwives or midwifery) **(0 hits)**
- (doctor* or clinician* or physician* or surgeon* or healthcare or health care or NHS or national health service*) **(100 hits)**
- GP* or general practitioner* or general practice or primary care) (7 **hits)**
- (dentist* or dental*). **(3 hits)**
- (pharmacist* or pharmacy).**(3 hits)**
- “occupational therap*” **(8 hits)**
- AHP* or “allied health profession*” or “allied health workforce” or PAMs or “professions allied to medicine”) **(158 hits)**

**^c^ Details of Cinahl searches from 2012 to current and limited to reviews (conducted 24^th^, 31^st^ January 2022)**

*Cinahl searches within titles and abstract for attract, recruit and retain*

- (recruit* or retain* or retention or attract*)
- (nurs*) **(327 hits)**
- (midwives or midwifery) **(13 hits)**
- (doctor* or clinician* or physician* or surgeon* or healthcare or “health care” or NHS or “National Health Service*”)) **(261 hits)**
- (GP* or “general practitioner” or “general practice” or “primary care”) **(72 hits)**
- (dentist* or dental) **(36 hits)**
- (pharmacist* or pharmacy) **(24 hits)**
- (occupational therap*) **(6 hits)**
- (physiotherap* or “physical therap*”) **(21 hits)**
- (radiographer* or radiologist*) **(0 hits)**
- (“speech and language therap*”) **(0 hits)**
- (“practitioner psychologist*” or “registered psychologist*”) **(0 hits)**
- (chiropodist* or podiatrist*) **(0 hits)**
- (paramedic*) **(12 hits)**
- (osteopath*) **(0 hits)**
- (orthotist*) **(0 hits)**
- (orthoptist*) **(0 hits)**
- (“operating department practi* or ODP*) **(0 hits)**
- (dietician* or dietetics) **(0 hits)**
- (“clinical scientist*” or “biomedical scientist*”) **(0 hits)**
- (“art therap*” or “music therap*” or “drama therap*”) **(0 hits)**
- (AHP* or “allied health profession*” or “allied health workforce” or PAMs or “professions allied to medicine”) **(10 hits)**

*Cinahl searches within titles and abstracts for return to practice*

- (return or re-ent* or reent* or re-licen* or relicen* or re-activate or reactivate or re-validate or revalidate or re-employ* or reemploy*) N10
- (nurs*) **(24 hits)**
- (midwives or midwifery) **(13 hits)**
- (doctor* or clinician* or physician* or surgeon* or healthcare or “health care” or NHS or “National Health Service*”) **(86 hits)**
- (GP* or “general practitioner” or “general practice” or “primary care”) **(8 hits)**
- (dentist* or dental) **(4 hits)**
- (pharmacist* or pharmacy) **(5 hits)**

## 9.2. Appendix 2: Summary table of reviews for return to practice

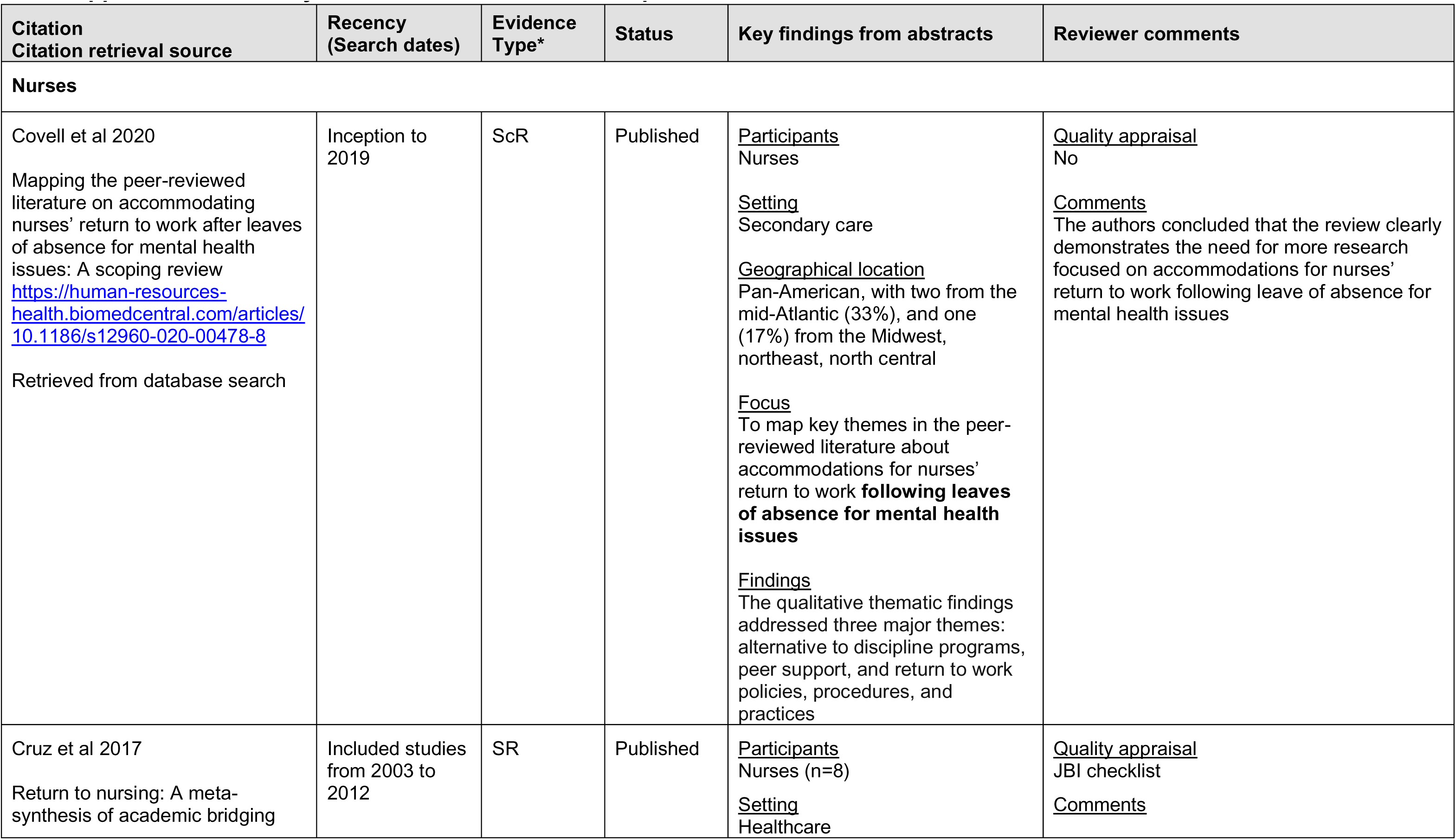
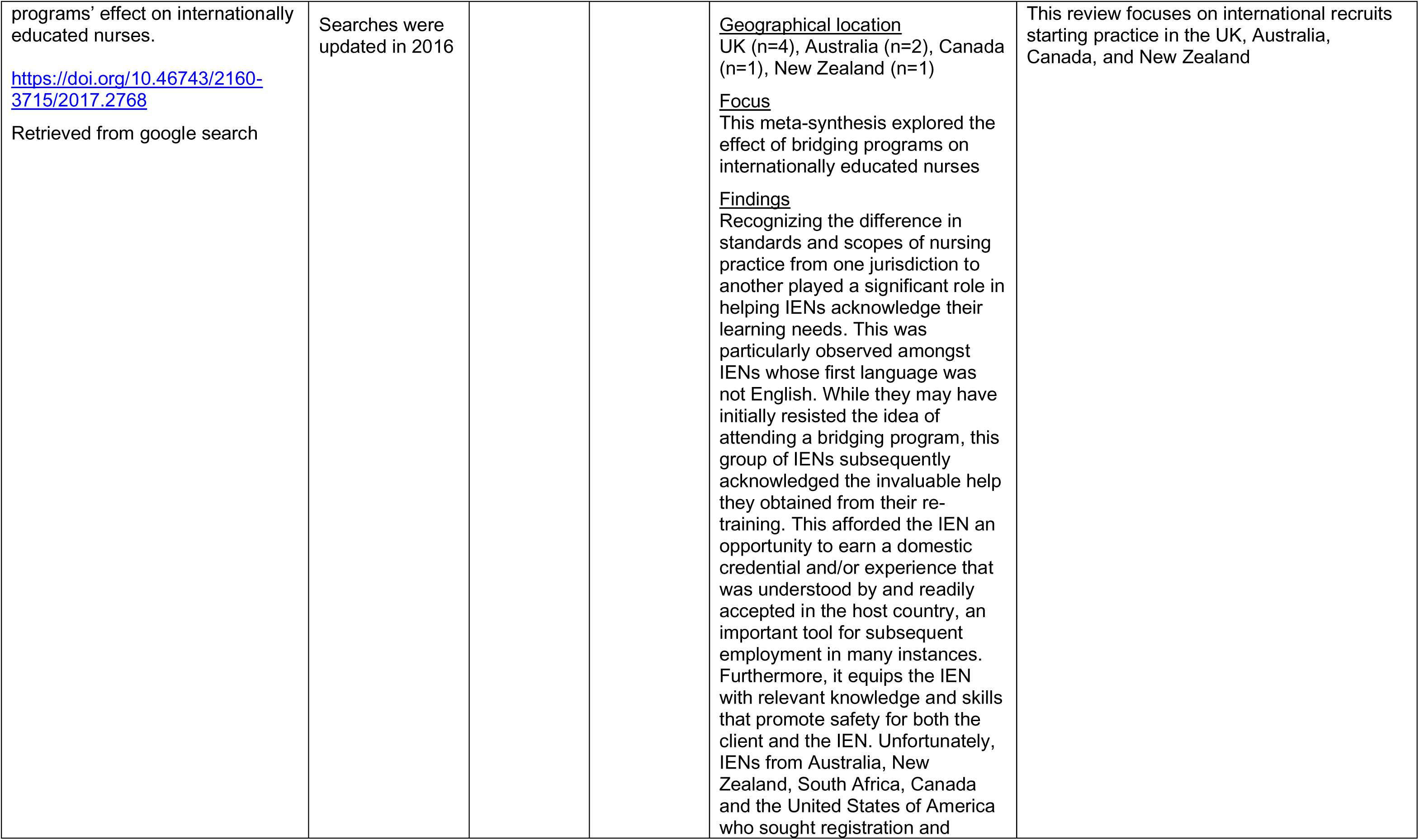
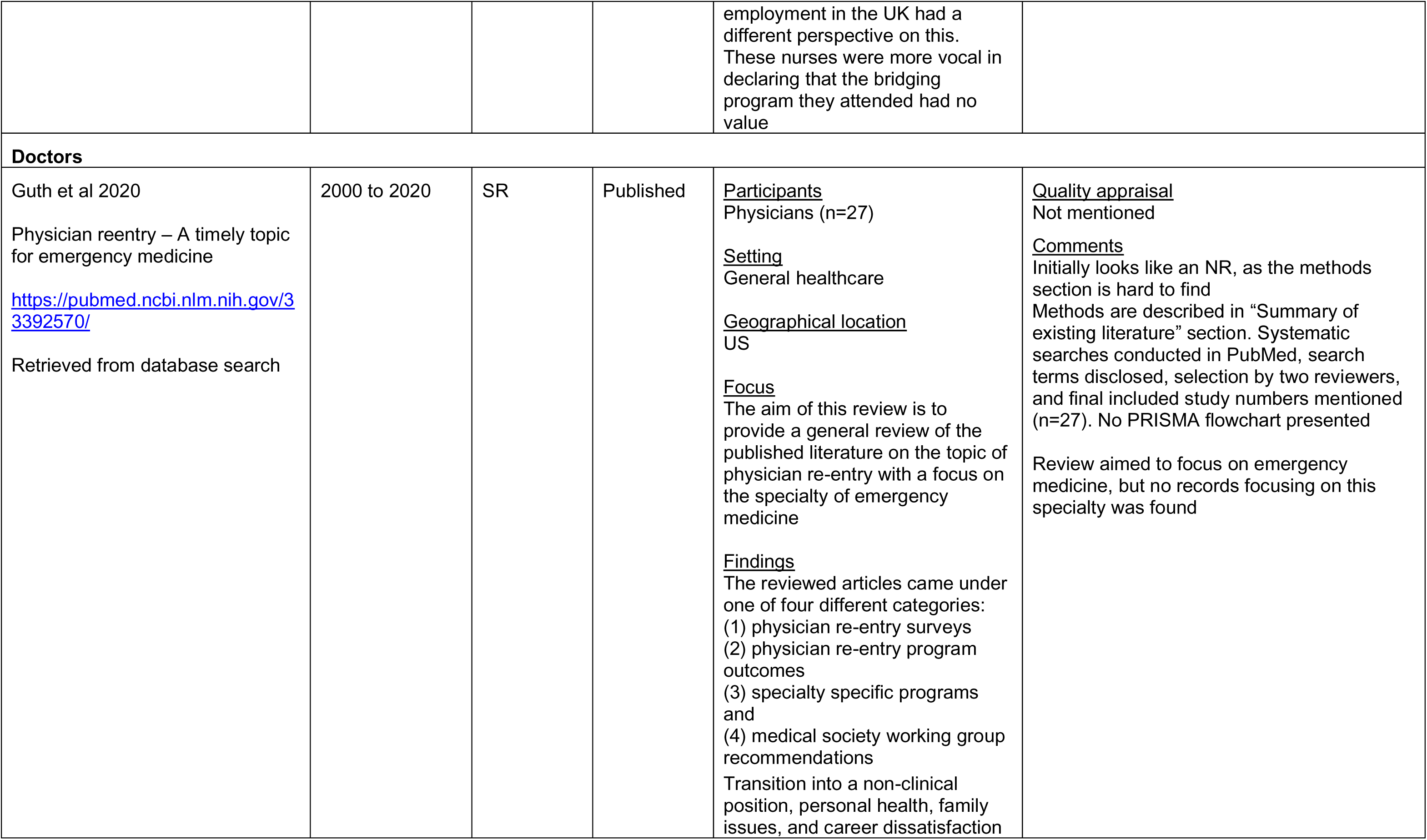
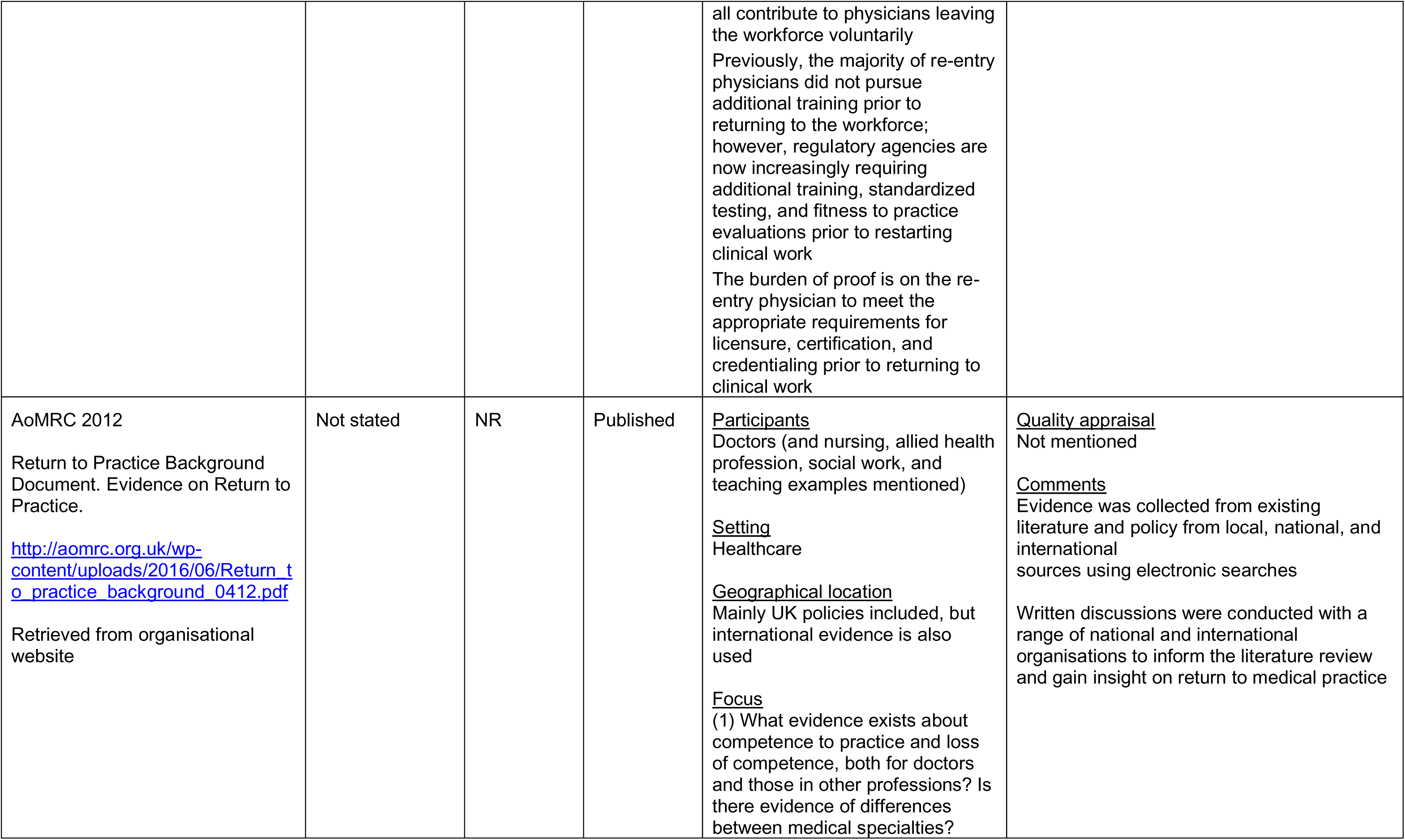
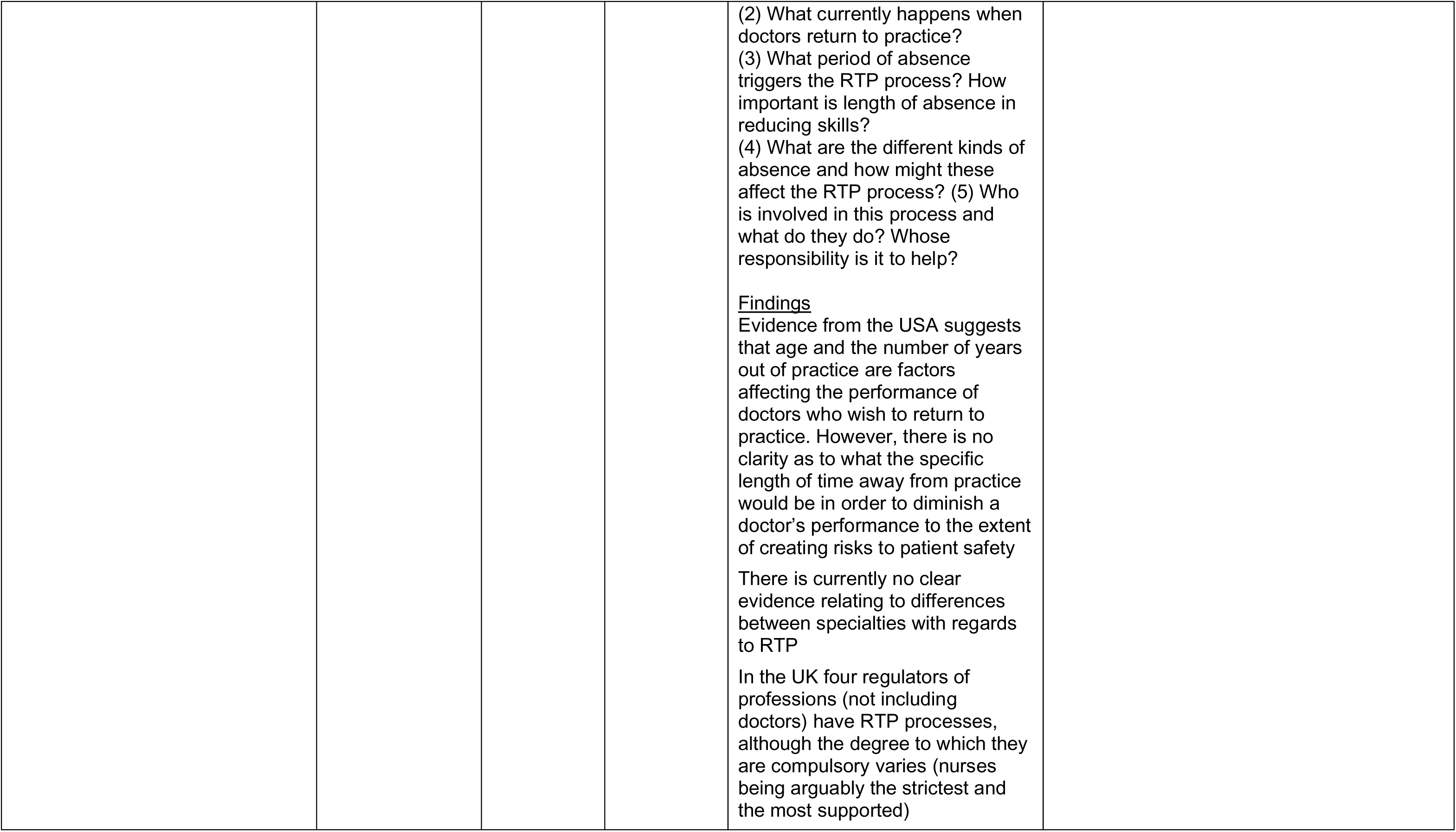
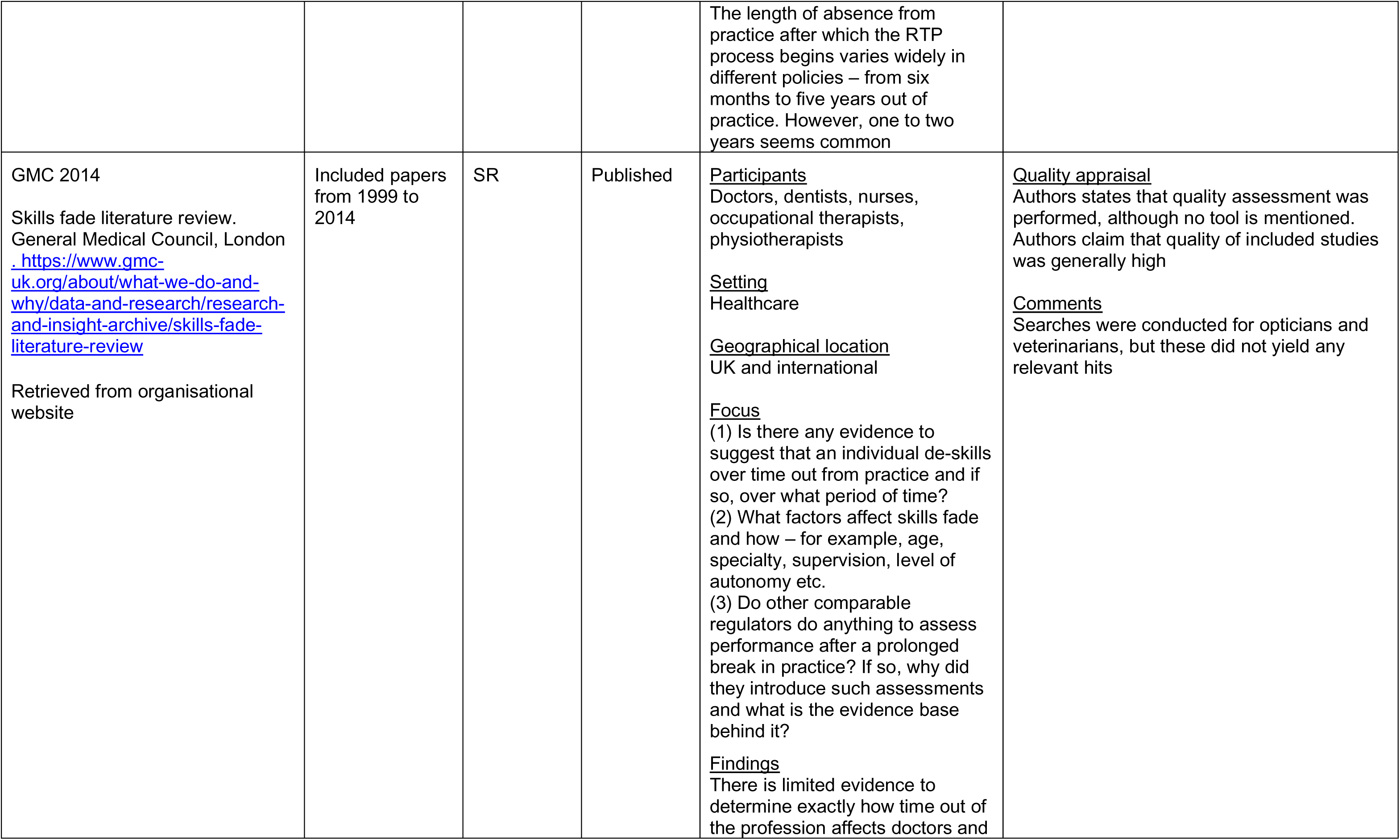
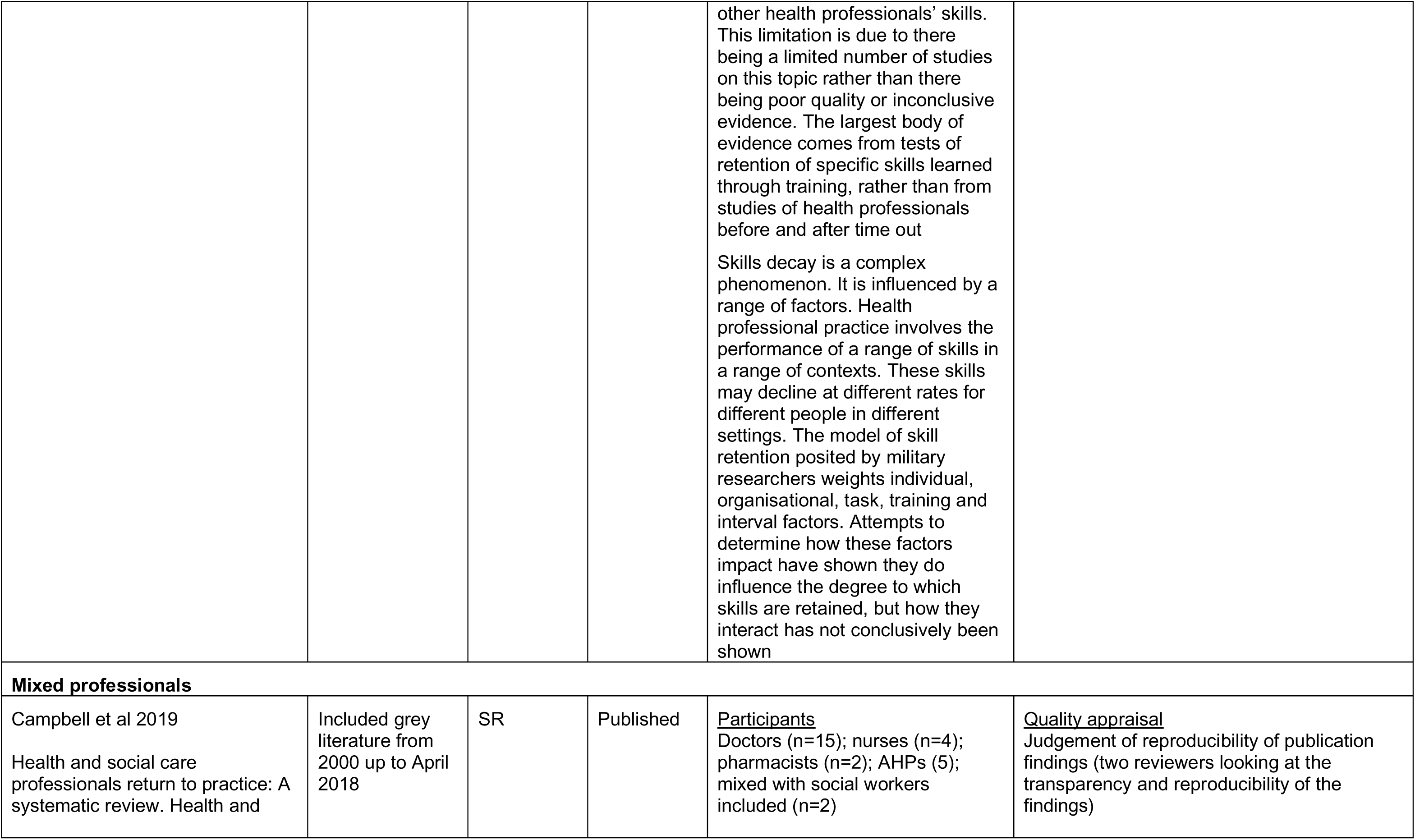
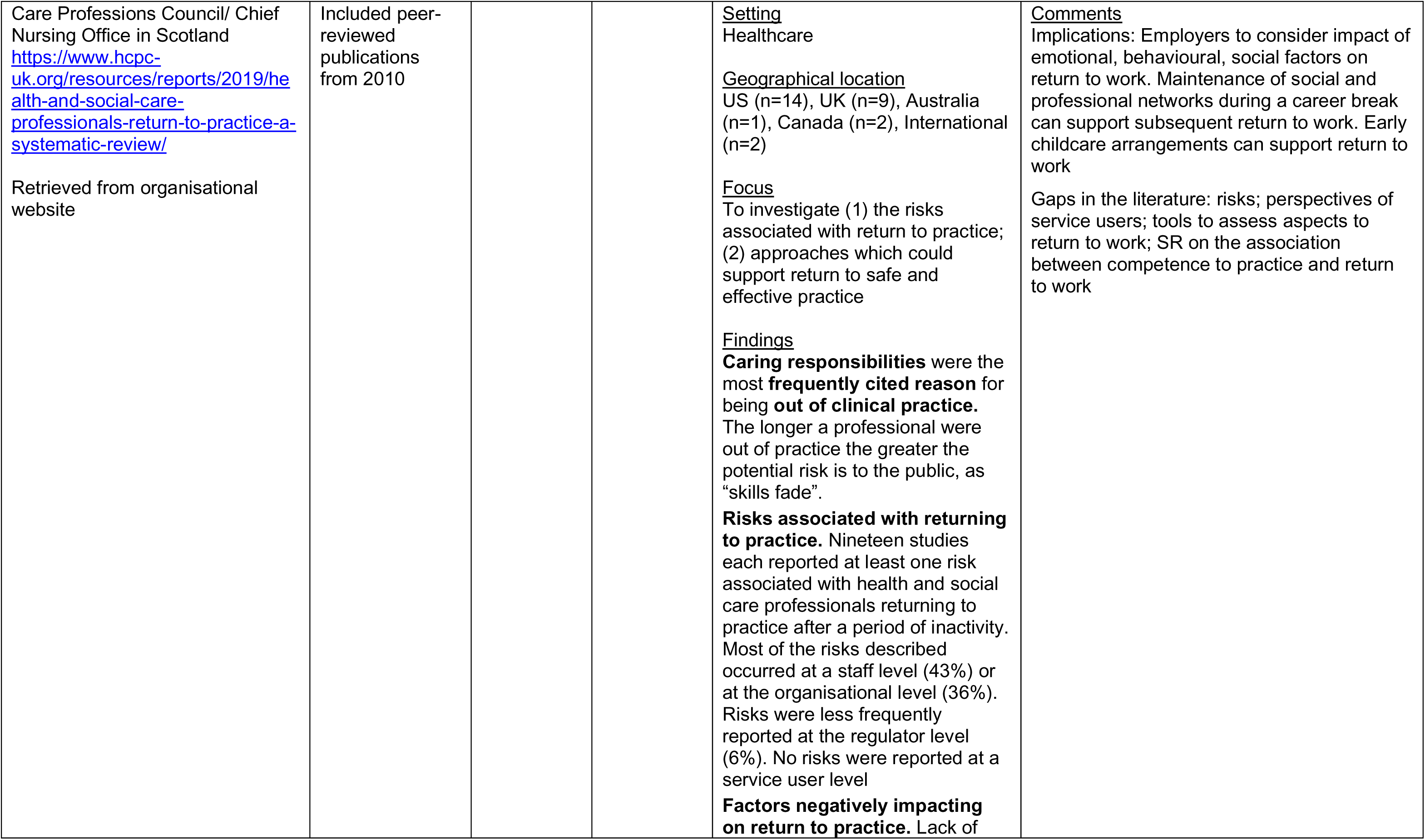
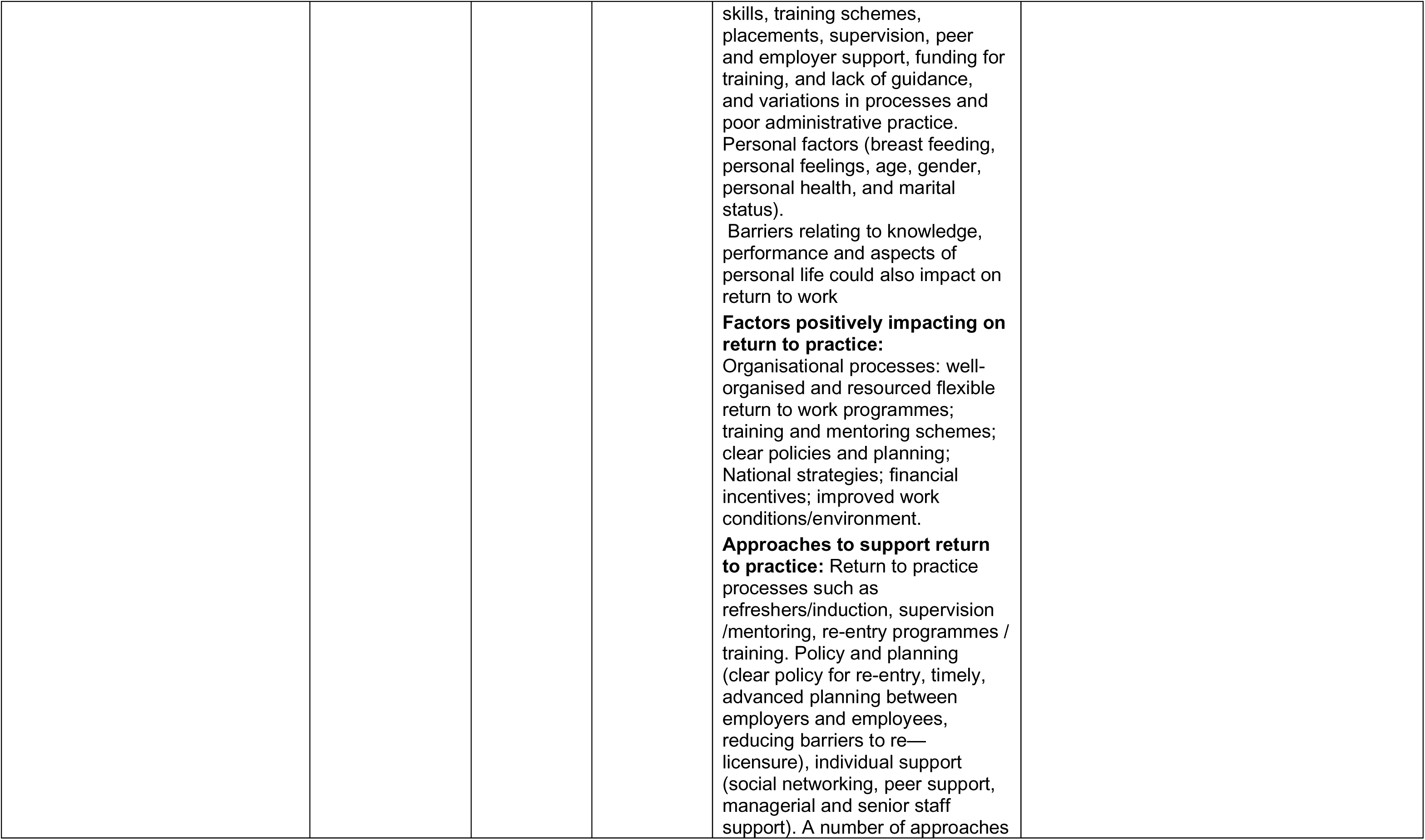
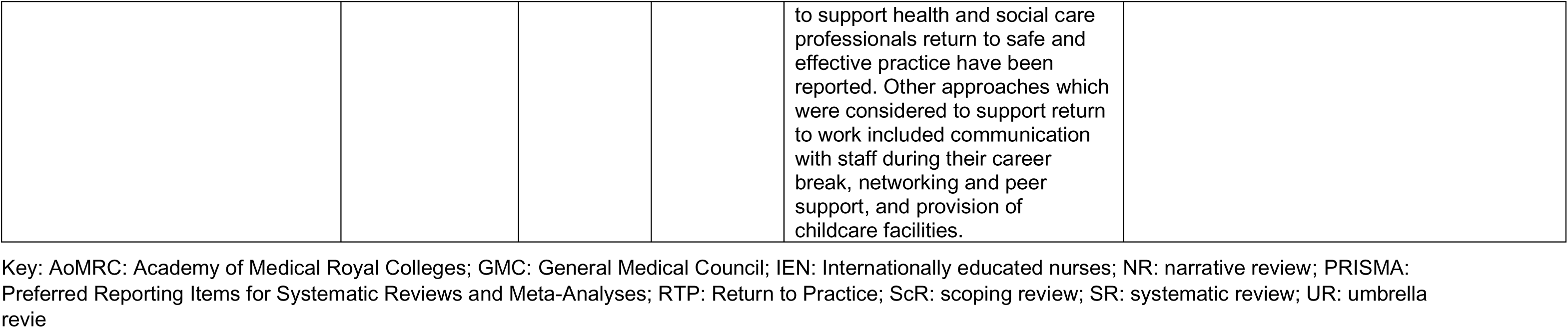

## 9.3. Appendix 3: Summary table of organisational reports for return to practice

| Citation<br>Citation retrieval source | Details | Key relevant findings and/or recommendations |
| --- | --- | --- |
| NHS England 2020<br>We are the NHS: People Plan 2020/21 – action for us all<br>www.england.nhs.uk/ournhspeople<br>Retrieved from organisational website | Report<br><u>Focus</u><br>This document sets out what the people of the NHS can expect – from their leaders and from each other – for the rest of 2020 and into 2021<br>This plan sets out actions to support transformation across the whole NHS. It focuses on how we must all continue to look after each other and foster a culture of inclusion and belonging, as well as action to grow our workforce, train our people, and work together differently to deliver patient care | Local recruitment (page 43)<br>Increasing local recruitment to roles such as clinical support workers<br>Growing apprenticeships<br>Expanding the primary care workforce<br>International recruitment (page 43)<br>Building local hubs<br>Increasing international recruitment<br>English language training<br>Co-ordinated international marketing<br>Health and care visa<br>Return to practice (page 44)<br>Encouraging former staff to return to the NHS<br>Supporting return to practice<br>Retaining our people (page 46)<br>Varied roles<br>Retaining people approaching retirement<br>Facilitating opportunities to retire and return<br>Retaining people in primary care<br>Support for retention |
| Health Education England 2014b<br>Nursing Return to Practice: Review of the current landscape. April 2014.<br>Health Education England, London.<br>https://www.hee.nhs.uk/sites/default/files/documents/HEE%20Return%20to%20practice%20-%20Review%20of%20the%20Landscape%20Apr%202014.pdf<br>Retrieved from organisational website | Report<br><u>Focus</u><br>The aim of this report was<br>(1) to identify the current nursing Return to Practice landscape<br>(2) to identify what works well and the challenges<br>(3) to scope what the opportunities are for the future. | Return to Practice (RTP) courses have low attrition rates and returning nurses to the workforce is far more cost effective than training a pre-registration nurse. However, the approach of RTP should be viewed as one option in a comprehensive strategy to increase the available workforce<br>There are some challenges in accessing RTP which focus upon variations in delivery in the UK and engagement of stakeholders:<br>Accessing information on RTP (e.g. on where and how to apply)<br>Having a clear local contact for RTP<br>The availability of supportive clinical placements and whether returners have to find their own placement<br>The capacity and quality of sign-off mentors<br>How supportive clinical areas are to returners |
|  |  | • Funding for students and trusts (some returners are funded, others self-fund and pay anything between £650 and £1500 in course fees and may have to find their own clinical placement)<br>• A joint trust and Higher Education Institution (HEI) approach to RTP<br>There is also a specific gap in RTP options in the community – most RTP nurses return to the acute sector initially, with very small numbers going into the community, mental health and children's branches<br>Developing a consistent approach to RTP will require changes in the commissioning and delivery of RTP in many regions, and a shift in the engagement of providers of clinical placements - all of which will require leadership and close working from Health Education organisations, providers and HEIs |

Key: HEI: Higher Education Institution; LTFT: Less Than Full-time Training; NHS: National Health Service; RTP: Return to Practice

## 9.4. Appendix 4: Summary table of primary research studies for return to practice

| Primary Research |  |  |  |  |  |
| --- | --- | --- | --- | --- | --- |
| Citation | Country | Type of study | Focus |  | Comments |
| **Nurses** |  |  |  |  |  |
| Barlow et al 2019 | UK | Mixed methods | Evaluation of a nursing RTP programme | Return to practice | Nurses on City University London RTP programme |
| Ipsos Mori 2016 | England | Qualitative | Views | Return to practice | Primary care nurses |
| Kenward et al 2017 | UK | Qualitative | Issues related to clinical contact time and to return to practice | Return to practice | Military nurses<br>From Campbell et al 2019<sup>a</sup> |
| McMurtrie et al 2014 | Australia | Quantitative | Satisfaction | Return to practice | Queensland Health Refresher Program |
| Noorland et al 2021 | Netherlands | Qualitative | Experiences and needs | Return to practice | During COVID-19 |
| **Doctors** |  |  |  |  |  |
| HEE 2020 | UK | Mixed methods | Evaluation of SuppoRTT | Return to practice | 1 year and 2 year evaluations |
| MacCuish et al 2021 | UK | Mixed methods | Evaluation of simulation training | Return to practice | Internal medicine |
| Mullender et al 2021 | UK | Quantitative | Evaluation of Springboard | Return to training | Physicians |
| RCOA 2021a | UK | Mixed methods | Factors influencing intention to stay longer or return after retiring | Retention<br>Return to practice | During COVID-19 |
| Randive et al 2015 | UK | Quantitative | Lengths and patterns of full-time and LTFT training | LTFT | Trainee anaesthetist's |
| Harries et al 2016 | UK | Quantitative | Evaluating the accessibility and experiences of flexible training | LTFT | Trainee surgeons |
| Green et al 2019 | USA | Quantitative | A case series in anaesthesiologist re-entry | Return to practice | Anaesthesiologists' re-entry program |
| **AHPs** |  |  |  |  |  |
| Thorn et al 2019 | UK | Mixed methods | Evaluation | Return to practice | AHPs and health care scientists |
| Dodds and Herkt 2013 | New Zealand | Qualitative | Experiences and management of return to practice after a career break | Return to practice | Occupational therapists |
| Hulcombe et al 2020 | Australia | Mixed methods | Describe flexible working arrangement, specifically part-time working for AHPs returning from maternity leave | Return to practice | AHPs |
| Parcsi and Curtin 2013 | Australia | Qualitative | Experiences of returning to work after maternity leave | Return to practice | Occupational therapists |
| Phipps et al 2013 | UK | Qualitative | Experiences of returning to work after career breaks or moving sectors | Return to practice | Pharmacists<br>From Campbell et al 2019<sup>a</sup> |
| **Mixed professional groups** |  |  |  |  |  |
| Coates and Macfadyen 2021 | UK | Qualitative | Experiences | Return to practice | Nurses, Midwives, AHPs |

<sup>a</sup> Secondary sourced from Campbell et al (2019). More details about Campbell et al. (2019) can be found in Table 3.
Key: AHPs: allied health professionals; LTFT: Less Than Full Time

## 9.5. Appendix 5: Mapping table for reviews of factors

| Citation | Type of review | Key Findings |
| --- | --- | --- |
| **Nurses** |  |  |
| Adams et al 2021<br>SR<br>23 included studies | <u>Focus</u>: Factors influencing retention<br><u>Population</u>: Nurses<br><u>Context</u>: Mental health services | Four key themes emerged from the synthesis: Individual characteristics, Working within mental health services, Training and skills and Work environment. The findings from this review suggest that MHNs encounter some factors unique to working in mental health services, which suggests that retention strategies should be specific to each nursing speciality. Beyond nursing speciality, the factors identified vary between clinical settings in mental health due to the differences in work environments and services they provide |
| Al Zamel et al 2020<br>SR<br>37 included studies | <u>Focus</u>: Factors influencing nurses' intention to leave and intention to stay<br><u>Population</u>: Nurses<br><u>Context</u>: Any setting | Many factors including job satisfaction, organizational commitment, quality of work life, work environment, leadership style, bullying at work, family reason, and job security were identified to be associated negatively with nurse's intention to leave and positively with intention to stay in organisation |
| Blay and Smith 2020<br>SR<br>11 included studies | <u>Focus</u>: Factors influencing recruitment and retention<br><u>Population</u>: Enrolled nurses<br><u>Context</u>: Any setting | Three major themes (Nursing work and the EN role, educational structure and support, and Personal attributes) were identified that covered enablers and barriers to the recruitment of Diploma of Nursing students and Enrolled Nurses. Evidence of the efficacy of programs designed to integrate and retain Enrolled Nurses in the health workforce is scant. Enrolled Nurses viewed participation in a Transition to Practice Program as an integral step in the pathway to becoming a registered nurse |
| Bloxsom et al 2019<br>SR<br>8 included studies | <u>Focus</u>: Factors associated with job satisfaction and intention to stay<br><u>Population</u>: Midwives<br><u>Context</u>: All maternity services | The data synthesised for this review clearly suggest that when midwives have good working relationships, are well supported by their managers, are able to develop relationships with the women in their care and can work in a normal birth centric model that offers variety and the opportunity to practise to the full scope of their role, they are inclined to stay in their jobs. Further, being able to practise their "passion" seemingly helps midwives get through the inevitable "rough days" |
| Brown et al 2013<br>SR<br>13 included studies | <u>Focus</u>: Factors influencing retention<br><u>Population</u>: Nurse managers<br><u>Context</u>: Front-line nurse or patient-care managers | Twenty-one factors were categorized into three major categories: organizational, role and personal. Job satisfaction, organizational commitment, organizational culture and values, feelings of being valued and lack of time to complete tasks leading to work/life imbalance, were prominent across all categories |
| Chamanga et al 2020<br>SR<br>10 included studies | <u>Focus</u>: Factors influencing retention<br><u>Population</u>: Adult nursing<br><u>Context</u>: Community settings | Data synthesis and analysis revealed individual and organisational factors influencing the retention of community nurses with the following three dominant themes: (1) work pressure, (2) working conditions and (3) lack of appreciation by managers |
| Falatah 2021<br>SR<br>43 included studies | <u>Focus:</u> Impact of the coronavirus disease (COVID-19) pandemic on turnover intention<br><u>Population:</u> Nurses<br><u>Context:</u> Any setting (10 COVID-19 studies) | The reviewed literature suggested that nurses' turnover intention increased significantly after the COVID-19 pandemic. Post-COVID-19-pandemic studies focused more on predicting nurses' turnover intention through the pandemic's negative impact on the nurses' psychological wellbeing |
| Hutchinson et al 2012<br>NR<br>Number of studies not reported in the abstract | <u>Focus:</u> Factors influencing attraction and retention<br><u>Population:</u> Nurses<br><u>Context:</u> Any setting | The literature review discussed the integration of the Y generation (born between 1980 and 2000) into the workforce and the need for the Health Service to recognise and prepare the current workforce to the Y Gen needs. They report an abundance of descriptions of the Y Gen, characteristics and values. They conclude that the focus should be on their strengths, and developments made to structure a workforce that supports the Y Gen in their professional nursing role. This needs to include an understanding of what attracts and retains the Y Gen although the authors recognise that there is limited reference to this in the literature reviewed |
| Hussain et al 2012<br>NR<br>Number of studies not reported in the abstract | <u>Focus:</u> Strategies for dealing with future shortages<br><u>Population:</u> Nursing workforce<br><u>Context:</u> All nurses in USA | This paper examines the intensity of the nursing shortage that exists in the health-care industry and evaluates the causes of this shortage. Factors responsible for the exodus of nurses from the profession include unpleasant workplace environment, poor communication within the organization, overwhelming stress, physical and verbal abuse, and personal health |
| Keith et al 2021<br>SR<br>Number of studies not reported in the abstract | <u>Focus:</u> Factors that influence intention to stay<br><u>Population:</u> Nurses<br><u>Context:</u> Millennial generation born 1981 – 1996 | The results from this review showed that Millennial generation nurses expect strong leadership, advancement opportunities, alignment of organizational and personal values, good co-worker relationships, healthy work-life balance, recognition and cutting edge technology. The authors concluded that millennials have specific expectations for work, and they will leave if these go unmet |
| Khan et al 2019<br>SR<br>15 included studies | <u>Focus:</u> Factors influencing intentions to leave<br><u>Population:</u> Adult nurses<br><u>Context:</u> Critical care | Three main themes emerged following data analysis. These themes were quality of the work environment, nature of working relationships and traumatic/stressful workplace experiences |
| MacKay et al 2021<br>SR<br>40 included studies | <u>Focus:</u> Influences on decisions to work in rural and remote healthcare settings<br><u>Population:</u> Nurses<br><u>Context:</u> Rural and remote healthcare settings | Thematic analysis identified three interrelated dimensions that influenced nurses' retention and migration decision-making: 'person/al', 'profession/al' and 'place' with 18 inter-related domains<br>The 'person/al' dimension contained five domains: a sense of belonging/connectedness, knowledge of rural culture, blurring of personal and professional lives, anonymity and job satisfaction/stress |
|  |  | The 'profession/al' dimension contained eight domains: expert generalist, advanced nurse practitioner, professional isolation, mentorship, education, autonomy and empowerment, role conflict, and recruitment and retention<br>The 'place' dimension contained five domains: terrain and weather, fewer resources, geographical isolation, safety and rural culture<br>The data informed the development of the MacKay's 3P (person/al, profession/al and place) model to capture the complex phenomenon of the influences on nurses' decision making to work in rural and remote settings |
| Marufu et al 2021<br>SR<br>46 included studies | <u>Focus:</u> Factors influencing recruitment and/or retention<br><u>Population:</u> Nurses<br><u>Context:</u> Any setting | Nine domains influencing staff turnover were found: nursing leadership and management, education and career advancement, organisational (work) environment, staffing levels, professional issues, support at work, personal influences, demographic influences, and financial remuneration |
| McClain et al 2022<br>ScR<br>38 included studies | <u>Focus:</u> Factors influencing retention<br><u>Population:</u> Nurses<br><u>Context:</u> Millennial generation born between 1981 – 1996 | Findings of this review are grouped under 5 concepts: leadership, work environment, professional growth, professional fatigue and self actualization (with barriers and strategies identified for each)<br>The review concluded that the current evidence base shows that barriers to and strategies for millennial nurse retention commonly focus on the work environment and the relationships between nursing leadership and the bedside nurse<br>A preliminary scan of the evidence indicates that creating a healthy work environment, that is collaborative, fair, flexible, challenging and provides opportunities for growth may keep millennial nurses engaged<br>Having a nursing leadership that models these values and leads by example may help millennial nurses to feel safe and supported |
| Nei et al 2015<br>SR<br>106 included studies | Focus: Factors influencing retention<br>Population: Nurses<br>Context: Any setting | Supportive and communicative leadership, network centrality, and organisational commitment are the strongest predictors of voluntary turnover based on meta-analytic correlations<br>Additional variables identified that relate to nurse turnover intentions include: job strain, role tension, work-family conflict, job control, job complexity, reward/recognition, and team cohesion. Findings suggest that some factors, such as salary, are relatively less important in the prediction of turnover |
| **Doctors** |  |  |
| Andah et al 2021<br>RR<br>82 included articles | <u>Focus:</u> Factors related to motivation for leaving<br><u>Population:</u> Medical professionals | Thematic analysis identified four key themes: low morale, disconnect, unmanageable change, and lack of personal and professional support. The themes of mastery, membership, and meaning were substantially present within the dataset |
|  | <u>Context:</u> Primary and secondary care |  |
| Darbyshire et al 2021<br>ScR<br>18 included studies | <u>Focus:</u> Factors associated with retention<br><u>Population:</u> Doctors<br><u>Context:</u> Emergency Medicine | Multiple factors were identified as linked with retention, including perceptions about teamwork, excessive workloads, working conditions, errors, teaching and education, portfolio careers, physical and emotional strain, stress, burnout, debt, income, work-life balance and antisocial working patterns |
| Holloway et al 2020<br>SR<br>41 included studies | <u>Focus:</u> Factors governing retention<br><u>Population:</u> Doctors<br><u>Context:</u> Rural and undeveloped areas in high-income countries | Papers were scrutinised for relevance to established rural recruitment and retention strategies and the key themes identified were: rural background, rurally focused education and training, personal and professional circumstances, and integration with the community<br>The major barriers to rural recruitment are family-unit considerations for partners and children, concerns over isolation and a poor perception of rural practice. Strategies to retain rural doctors need a greater focus on personal and professional support networks and community integration |
| Mohammadiaghdam et al 2020<br>SR<br>35 included studies | <u>Focus:</u> Determining factors in retention<br><u>Population:</u> Physicians<br><u>Context:</u> Rural and underdeveloped areas | The major affecting factors in physicians' retention in rural and underdeveloped regions were classified into the following six categories: 1) financial; 2) career and professional; 3) working conditions; 4) personal; 5) cultural; and 6) living conditions factors<br>Working conditions factors and financial factors deserve healthcare policy makers' particular attention among the factors which are associated with the retention and willingness of physicians to serve in deprived areas<br>Recruiting physicians, who are from rural backgrounds and rural origins, is another determining factor in physicians' retention which has to be considered by the policy makers who aim to promote the physicians' retention in rural areas<br>There is not enough evidence regarding the cultural factors and their effect on the physicians' retention in the mentioned areas |
| **GPs** |  |  |
| Asghari et al 2020<br>UR<br>14 included reviews | <u>Focus:</u> Factors influencing recruitment and/or retention<br><u>Population:</u> Family physicians<br><u>Context:</u> Rural | Fourteen SRs met the inclusion criteria, from which 158 specific factors were identified and summarised into 11 categories: personal, health, family, training, practice, work, professional, pay, community, regional and system/legislation. The three categories referenced most often were training, personal and practice. The specific individual factors mentioned most often in the literature were 'medical school characteristics', 'longitudinal rural training' and 'raised in a small town'<br>The three most often cited categories resemble three distinct phases of a family physician's life: pre-medical school, medical school and post-medical school. To increase the number of physicians who choose to work in rural practice, strategies must encompass and promote continuity across all three of these phases |
| Mitchell et al 2018<br>Rapid ScR<br>50 included studies | <u>Focus:</u> Factors influencing recruitment, retention and returning to work<br><u>Population:</u> Primary care<br><u>Context:</u> Included papers were mainly from England and the UK, with some from Europe, USA, and Australia | A range of factors is leading to the decreasing number of GP recruits (e.g. perceptions of general practice as an unpopular medical career choice, a negative portrayal of general practice in medical schools and in society generally and a perception of less potential for career progression)<br>A range of factors is leading to GPs to leave the profession, including an unmanageable workload with poor support and constant organisational change, a perception that the profession not valued, a perceived lack of autonomy and support<br>Additionally, changes to work visas and regulatory requirements may affect GPs from abroad which may change according to government policies |
| Parlier et al 2018<br>NR<br>83 included studies | <u>Focus:</u> Factors that help develop, recruit, and retain<br><u>Population:</u> Primary care physicians<br><u>Context:</u> Rural - United States, Canada, or Australia | The authors' proposed a theoretical model that suggests factors interact across multiple dimensions to facilitate the development of a rural physician identity. Rural upbringing, personal attributes, positive rural exposure, preparation for rural life and medicine, partner receptivity to rural living, financial incentives, integration into rural communities, and good work-life balance influence recruitment and retention |
| Viscomi et al 2013<br>SR<br>86 included studies | <u>Focus:</u> Factors influencing attraction, recruitment, and retention<br><u>Population:</u> Primary care<br><u>Context:</u> Rural and remote areas of Australia and Canada | Factors such as rural background (of medical student or partner, or both), male gender, interest in living in a rural area and meaningful rural elective exposure during medical training were some of the important indicators related to rural practice |
| Wieland et al 2021<br>SR<br>6 included studies | <u>Focus:</u> Factors influencing retention<br><u>Population:</u><br><u>Context:</u> Remote areas of Australia and Canada | Six synthesised findings were identified: peer and professional support, organisational support, uniqueness of remote lifestyle and work, burnout and time off, personal family issues and cultural and gender issues<br>Long-term retention of doctors in remote areas of Australia and Canada is influenced by a range of negative and positive perceptions, and experiences with key factors being professional, organisational and personal |
| Ogden et al 2020<br>SR<br>27 included studies | <u>Focus:</u> Factors recruitment and retention<br><u>Population:</u> Undergraduates and postgraduates<br><u>Context:</u> Rural | GPs with rural backgrounds or rural experience during undergraduate or postgraduate medical training are more likely to practise in rural areas<br>The effects of multiple rural pipeline factors may be cumulative, and the duration of an experience influences the likelihood of a GP commencing and remaining in rural general practice |
| **AHPs** |  |  |
| Couch et al 2021<br>SR | <u>Focus:</u> Factors influencing recruitment and/or retention | Of the 22 included studies, 12 reported organisational/workplace structure and personal factors positively impacting recruitment and 11 studies discussed organisational and workplace structure also negatively impacting on retention |
| 22 included studies | <u>Population:</u> Allied Health Professionals<br><u>Context:</u> Metropolitan, rural, and remote locations | Career opportunities positively impacted on recruitment, while lack of opportunity negatively affected retention<br>Previous location exposure positively impacted recruitment however had limited impact on retention. Similarly, a diverse clinical load was reported as being attractive during recruitment, but unmanageable caseloads affected retention |
| Pretorius et al 2016<br>NR<br>24 included studies | <u>Focus:</u> Factors influencing retention<br><u>Population:</u> Physiotherapists<br><u>Context:</u> Australia | Strategies to improve the retention of skilled physiotherapists were broadly grouped into improving professional support in the workforce and assisting the re-entry process for physiotherapists seeking to return to the workforce. |
| Roots and Li 2013<br>SR<br>12 included studies | <u>Focus:</u> Factors influencing recruitment and/or retention<br><u>Population:</u> Occupational therapists and physiotherapists<br><u>Context:</u> Rural areas | The decision to locate, stay or leave rural communities was influenced by: availability of and access to practice supports, opportunities for professional growth and understanding the context of rural practice (such as larger caseloads, limited referral options, decreased access to resources and limited access to continuing education)<br>The second-order analysis revealed the benefits of a strength-based inquiry in determining recruitment and retention factors. The themes that emerged were 1) support from the organization influences retention, 2) with support, challenges can become rewards and assets, and 3) an understanding of the challenges associated with rural practice prior to arrival influences retention<br>The review concludes how universally important practice supports are in the recruitment and retention of rehabilitation professionals in rural practice |
| **Mixed groups of healthcare professionals** |  |  |
| Jamebozorgi et al 2021<br>SR<br>23 included studies | <u>Focus:</u> Effects of the Covid-19 pandemic on the employees and the retention strategies<br><u>Population:</u> Health workers<br><u>Context:</u> Any setting | Stigma and violence against the staff, burnout and stress, increased staff workload, acquisition of communication skills, employees' mental and physical health, employee safety during disaster, staff mobilization to assist the current forces, expansion of cyberspace infrastructures, and motivational-health incentives were selected as codes<br>The authors concluded that retention or non-retention of the personnel during or after a disaster can be caused by the multifaceted effects of the crisis on people. Thus, a combination of several appropriate strategies should be used to respond to it in order to reduce the adverse effects of the disasters |
| Koebisch et al 2020<br>SR<br>5 included studies | <u>Focus:</u> Factors that are most important in recruitment and retention<br><u>Population:</u> Chiropractors, osteopaths, dentists and physiotherapists. But only papers for physicians retrieved<br><u>Context:</u> Rural areas in Canada | Identified five themes in two domains, recruitment and retention: personal/family matters, community factors, professional practice factors, professional education factors and economic factors<br>Forty major codes were generated through axial coding of open codes. Codes included attraction to rural lifestyle, recreational activities, Scope of practice, rural training and incentives |
|  |  | Scope of practice was deemed very important as a factor of recruitment, as was attraction to rural lifestyle. Incentives were found to be of little importance in influencing the recruitment of healthcare professionals, and even less important for retention<br>A lack of research was determined in the realm of factors influencing the recruitment and retention in healthcare professionals other than medical doctors in Canada |
| Mbemba et al 2016<br>UR<br>15 included reviews | <u>Focus:</u> Factors Influencing recruitment and retention<br><u>Population:</u> Healthcare workers<br><u>Context:</u> Rural and remote areas in developed and developing countries | The most important factors influencing recruitment were rural background and rural origin, followed by career development<br>The main factors impacting retention were opportunities for professional advancement, professional support networks and financial incentives<br>While the main factors influencing recruitment and retention have been largely explored in the literature, the evidence on strategies to reduce the shortage of healthcare workers in rural area, particularly in developing countries, is low |
| RCOA 2021b<br>RR<br>140 included studies | <u>Focus:</u> Factors affecting retention<br><u>Population:</u> Anaesthetists, surgeons, and other NHS professionals (hospital nurses, GPs, paramedics, mental health staff)<br><u>Context:</u> Developed countries, metropolitan and rural. | The key factors identified as influencing whether anaesthetists and others stay in their roles:<br>*Individual-level factors:* mental wellbeing and burnout; physical issues associated with aging; the extent to which professionals felt valued and satisfied with their work; and family commitments and other priorities<br>*Role-related factors:* workload and working requirements, including working on call; plus perceived autonomy in the role<br>*Organisational / team-related factors:* organisational climate; leadership; communication; team morale; and supportive relationships<br>*System-level factors:* perceived bureaucracy; issues related to income and pensions; and concerns about litigation or risks<br>Although the above factors were identified the review states that it found little good evidence about the best ways to retain professionals in the NHS<br>Teams tested strategies such as peer support, reduced hours, bonuses and portfolio roles, however it is difficult to say whether these are effective<br>Some studies looked at professionals' intentions but did not follow up to see whether people stayed in their roles. Others found improvements in job satisfaction or wellbeing but did not see whether this encouraged people to keep working in the NHS |
| Russell et al 2017<br>SR<br>8 included studies | <u>Focus:</u> Determinants of retention<br><u>Population:</u> Primary health care workers (AHPs and GPs)<br><u>Context:</u> Rural Australia | The review recognises a broad range of factors that are associated with the rural retention of Australian primary health care workers including: geographical remoteness and population size, profession, providing hospital services, practising procedural skills, taking annual leave, employment grade, employment and payment structures, restricted access to provider numbers, country of training, vocational training, practitioner age group and cognitive behavioural coaching |
|  |  | They conclude that the findings suggest that retention strategies should be multifaceted and bundled |
| Terry et al 2021<br>SR<br>13 included studies | <u>Focus:</u> Factors influencing recruitment and/or retention<br><u>Population:</u> Pharmacists<br><u>Context:</u> Rural | Study-specific factors associated with recruitment and retention of pharmacists in rural practice were identified and grouped into five main themes: geographic and family-related, economic and resources, scope of practice or skills development, the practice environment, and community and practice support factors |

Key: GP: General Practitioner; MHN: mental health nurse, NHS: National Health Service; NR: narrative review; ScR: scoping review; RCOA: Royal College of Anaesthetists RR: rapid review; SR: systematic review; UR: umbrella review

## 9.6. Appendix 6: Mapping table for reviews of interventions/strategies

| Citation<br>Type of review<br>Extent of evidence | Study details<br>Focus / Population / Context | Key Findings |
| --- | --- | --- |
| **Nurses** |  |  |
| Halter et al 2017<br>UR<br>7 included reviews | <u>Focus:</u> Interventions to reduce turnover<br><u>Population:</u> Adult nurses<br><u>Context:</u> Hospital and community settings, mostly USA | Moderate quality review evidence, albeit from poorly controlled primary studies of a small number of interventions which decrease turnover or increase retention of nurses, these being preceptorship of new graduates and leadership for group cohesion |
| Mbemba et al 2013<br>UR<br>5 included reviews | <u>Focus:</u> Interventions for supporting retention<br><u>Population:</u> Nurses<br><u>Context:</u> Rural and remote areas | Two reviews showed that financial-incentive programs have substantial evidence to improve the distribution of human resources for health<br>The other three reviews highlighted supportive relationships in nursing (mentoring, clinical supervision and preceptorship), information and communication technologies support and rural health career pathways as factors influencing nurse retention in rural and remote areas |
| McClain et al 2022<br>ScR (no evaluation)<br>36 included studies | <u>Focus:</u> Summary of strategies influencing retention<br><u>Population:</u> Registered nurses born between 1980 and 2000<br><u>Context:</u> Any setting | The current evidence base shows that barriers to and strategies for millennial nurse retention commonly focus on the work environment and the relationships between nursing leadership and the bedside nurse<br>A preliminary scan of the evidence indicates that creating a healthy work environment that is collaborative, fair, flexible, challenging, and provides opportunities for growth may keep millennial nurses engaged |
| Redknap et al 2015<br>NR<br>Number of studies not reported | <u>Focus:</u> Examines the impact the nursing practice environment has on retention<br><u>Population:</u> General and mental health nurses<br><u>Context:</u> General and mental health settings | Findings indicate, that while there is a wealth of evidence to support the importance of a positive practice environment on nurse retention in the broader health system, there is little evidence specific to mental health<br>Further research of the mental health practice environment is required |
| Twigg and McCullough 2014<br>NR<br>39 included studies | <u>Focus:</u> Strategies to create and enhance positive practice environments<br><u>Population:</u> Nurses<br><u>Context:</u> Clinical settings. | Strategies for creating a positive practice environment included: empowering work environment, shared governance, structure, autonomy, professional development, leadership support, adequate numbers and skill mix and collegial relationships within the healthcare team<br>Creating positive practice environments enhances nurse retention and facilitates quality patient care. Managers and administrators should assess and manage their practice environments using a validated tool to guide and evaluate interventions |
| **Doctors** |  |  |
| Darbyshire et al 2021<br>ScR (no evaluation)<br>18 included studies | <u>Focus:</u> Effectiveness of interventions to improve retention<br><u>Population:</u> Doctors<br><u>Context:</u> Emergency Medicine | The majority of papers did not directly address efforts to improve retention<br>Of those that did, only a small number reported conclusions from empirical work<br>Studies mainly offered suggestions from a range of perspectives which included improving workflow and staffing<br>Self-care and compassion dialogues and work scheduling (rostering) |
| Kumar and Clancy 2020<br>SR<br>19 included studies | <u>Focus:</u> Effectiveness of interventions to improve retention<br><u>Population:</u> Physicians and surgeons<br><u>Context:</u> Rural areas | Financial incentives were less influential on retention, but results were inconsistent between studies and differed between high-, middle- and low-income nations<br>Successful strategies included student selection from rural backgrounds into medical school and undergraduate education programs and early postgraduate training in a rural environment<br>Bundled or multifaceted interventions may be more effective than single factor interventions |
| Mallett et al 2022<br>ScR (no evaluation)<br>16 included studies | <u>Focus:</u> Strategies to enhance recruitment and retention<br><u>Population:</u> Paediatricians<br><u>Context:</u> UK, US, Canada | There is a paucity of data in the literature that describes evidence-based approaches to enhancing retention and recruitment in paediatrics.<br>The most important strategies employed to help are identified and grouped into six main themes<br>These include professional advocacy, workforce diversity, mentorship, improving working conditions, career flexibility and enhancing educational opportunities |
| Thi Nguyen et al 2021<br>NR<br>Number of studies not reported | <u>Focus:</u> Developing guidelines for strategies to attract, recruit, retain<br><u>Population:</u> Less attractive specialities, such as generalists, doctors<br><u>Context:</u> In rural areas, preventative vs curative services | By reducing feelings of professional isolation and reinforcing feelings of competence and autonomy in physicians, continuing medical education activities show promise as a strategy to recruit and retain physicians in less attractive specialties |
| Johnson et al 2018<br>SR<br>62 included papers | <u>Focus:</u> Rural placement programs within medical education<br><u>Population:</u> Medical students<br><u>Context:</u> Rural clinical placement or program | Rural background is an important aspect for program planners to consider, however our review identifies there are other key potential rural predictors including; rural interest/intentions prior to the program, generalist practice intentions, an interest in primary care and family medicine, financial and rural bonded scholarships and importantly the type and quality of a rural immersion experience and its duration |
| **GPs** |  |  |
| Marchand and Peckham 2017<br>Peckham et al 2016<br>SR<br>26 included studies | <u>Focus:</u> Approaches to retention and recruitment<br><u>Population:</u> GPs and family doctors<br><u>Context:</u> Countries that are part of the OECD | Studies that examine specific recruitment strategies for the GP workforce are scarce. This review suggests that most studies on primary care physician recruitment (for example, GPs and family doctors) have predominantly focused on remote rural locations |
| Mitchell et al 2018<br>Rapid ScR (no evaluation)<br>50 included studies | <u>Focus:</u> Strategies relating to recruitment retention and returning to work<br><u>Population:</u> Primary Care<br><u>Context:</u> Mostly carried out in the USA, Canada and Australia, particularly in rural locations | A number of strategies to improve early orientation towards GP recruitment has been suggested, (e.g. improved funding for clinical placements, encouragement of respect between medical professionals, inspiring GP role models and leaders, improving the public image of general practice through outreach work in schools and with the public)<br>Strategies to improve retention relate to trying to increase capacity and reduce workload, encourage variation in working life through portfolio careers and sub-specialisms as well as greater support for those wishing to change their clinical workload |
| Verma et al 2016<br>SR<br>51 included studies | <u>Focus:</u> Strategies to recruit and retain<br><u>Population:</u> Primary Care<br><u>Context:</u> International | A review of 51 studies included 42 different interventions to recruit or retain GPs<br>Retention initiatives mainly focused on financial incentives, wellbeing or peer support initiatives and support for professional development or research<br>There was limited evidence of effectiveness about most interventions and mixed evidence about financial rewards |
| **Dentists** |  |  |
| Suphanchaimat et al 2016<br>SR<br>7 included studies | <u>Focus:</u> Rural-exposure strategies on the intention to practice<br><u>Population:</u> Dental students and dental graduates<br><u>Context:</u> Rural areas | Enrolling students with rural backgrounds and imposing compulsory clinical rotation in rural areas during their study appeared to be effective strategies in tackling the shortage and maldistribution of dentists in rural area |
| **AHPs** |  |  |
| Obamiro et al 2020<br>ScR (no evaluation)<br>13 included studies | <u>Focus:</u> Strategies to increase the workforce<br><u>Population:</u> Pharmacists<br><u>Context:</u> Rural and remote in Australia | Enrolment of students from rural backgrounds, availability of support personnel for rural initiatives, extended rural placement and the inclusion of rural content in the teaching curriculum |
| Pretorius et al 2016<br>NR<br>24 included studies | <u>Focus:</u> Strategies influencing retention<br><u>Population:</u> Physiotherapists<br><u>Context:</u> Australia | Strategies to improve retention of skilled physiotherapists were broadly grouped into improving professional support in the workforce and assisting the re-entry process for physiotherapists seeking to return to the workforce |
|  |  | Many physiotherapists in Australia leave the workforce and the profession early in their careers. Addressing modifiable factors of attrition could help improve the retention of practitioners and skills in the profession, building workforce capacity |
| **Mixed groups of healthcare professionals** |  |  |
| Esu et al 2021<br>UR<br>9 included systematic reviews | <u>Focus:</u> Interventions for improving attraction and retention<br><u>Population:</u> Health workers<br><u>Context:</u> Rural and underserved areas | Recruiting rural students and rural placements improved attraction and retention although most studies were without control groups, which made conclusions on effectiveness difficult |
| Grobler et al 2015<br>SR<br>1 included study | <u>Focus:</u> Interventions to increase the rural health workforce<br><u>Population:</u> Health professionals<br><u>Context:</u> Rural | This review found one new study that evaluated the effect of a National Health Insurance scheme on the distribution of health professionals in Taiwan<br>The implementation of a National Health Insurance scheme made medical care more affordable for all Taiwanese citizens in both urban and rural areas<br>This may have led to better geographical distribution of health professionals (low certainty) |
| HEE 2014a<br>NR<br>20 included papers | <u>Focus:</u> Evaluation of values-based recruitment<br><u>Population:</u> Employees in NHS<br><u>Context:</u> UK | The research literature identified in this review provides several important insights regarding the impact of value congruence between employees and organisations that are highly relevant to the implementation of VBR in healthcare<br>However, the organisational (contextual) differences in this literature indicate that results should be interpreted with some caution as outcomes may not be immediately generalisable to a healthcare context and in particular to the NHS<br>Whilst the drivers for implementing VBR into the NHS are focused around the need to ensure the best possible care for patients, consistently, across professional, institutional and geographical boundaries, the literature on employees' values in other contexts and occupations may have different drivers, for example, to improve job satisfaction and productivity, or reduce staff turnover |
| Jelyani et al 2021<br>ScR (no evaluation)<br>50 included studies | <u>Focus:</u> Interventions for improving retention<br><u>Population:</u> Healthcare workers (all health workers (n=39); physicians (n=5); nurses (n=5); dentists (n=1))<br><u>Context:</u> During epidemics | According to the analysis, the interventions were classified into five themes (preparation, protection, support, care and, feedback) and 15 sub-themes (creating communication networks, providing education programs, telemedicine, protective measures, and equipment supply, revising and adjusting work shifts, early detection, organizational psychological |
|  |  | support, peer support, welfare support, professional support, smart human resource utilization, providing psychological care services, providing non-psychological care services, getting active feedback, and getting passive feedback)<br>Finally, it seems that to strengthen and sustain human resources in the face of epidemics, we must pay attention to various dimensions<br>Therefore, taking action in each of these themes cannot be helpful independently. It is recommended that managers and decision-makers implement strategies that cover more themes and are adjusted to their context |
| Patterson et al 2016<br>NR<br>20 studies included | <u>Focus:</u> Impact of value congruence on outcomes<br><u>Population:</u> Education providers/ students, trainers/ trainees and employers/employees<br><u>Context:</u> Recruiting for values | Practical implications are discussed in the context of values-based recruitment in the healthcare context<br>Theoretical implications of our findings imply that prosocial implicit trait policies, which could be measured by selection tools such as situational judgment tests, structured interviews and multiple-mini interviews, may be linked to individuals' values via the behaviours individuals consider to be effective in given situations<br>Further research is required to state this conclusively however, and methods for values-based recruitment represent an exciting and relatively uncharted territory for further research |
| RCOA 2021b<br>RR<br>10 included studies (anaesthetists and surgeons)<br>42 included studies and 6 systematic reviews (other NHS professionals) | <u>Focus:</u> Strategies for improving retention<br><u>Population:</u> Anaesthetists, surgeons, and other NHS professionals (hospital nurses, GPs, paramedics, mental health staff)<br><u>Context:</u> UK and international literature | The review identified few studies about strategies to improve the retention of anaesthetists or surgeons<br>The studies available tended to focus on improving mental wellbeing or job satisfaction as a proxy for potentially increasing retention<br>For other NHS professionals there were a number of interventions aiming to improve retention which included: Support initiatives, professional development, reimbursement and terms, other initiatives<br>Most of the research suggested positive short-term outcomes at the level of the individual, such as reduced stress or burnout.<br>Some found that staff were more likely to say they intended to stay in their roles<br>Most studies did not explore the medium to longer term impact on retention (whether staff actually did remain in their roles) and focused largely on individual or small team factors, rather than broader organisational or system-level approaches |
|  |  | It was noted that interventions targeting multiple levels may be more advantageous |
| Kroezen et al 2015<br>NR<br>64 included studies | <u>Focus:</u> Recruitment and retention strategies<br><u>Population:</u> Health professionals<br><u>Context:</u> Europe | The reviewers identified 64 studies about 34 different interventions in 20 European countries, including the UK<br>However, there was a lack of evidence about whether the strategies were effective, and most interventions were not part of a coherent package of approaches<br>The reviewers suggested single R&R interventions on their own have little impact, bundles of interventions are more effective<br>Interventions backed by political and executive commitment benefit from a strong support base and involvement of relevant stakeholders |

Key: GP: General Practitioner; MMIs: multiple mini-interviews, NHS: National Health Service; NR: narrative review; ScR: scoping review; RCOA: Royal College of Anaesthetists RR: rapid review; SJTs: situational judgement tests, SR: systematic review; UR: umbrella review

